# Population Impact and Efficiency of Improvements to HIV PrEP Under Conditions of High ART Coverage among San Francisco Men Who Have Sex with Men

**DOI:** 10.1101/2021.03.11.21253406

**Authors:** Adrien Le Guillou, Susan Buchbinder, Hyman Scott, Albert Liu, Diane Havlir, Susan Scheer, Samuel M. Jenness

**Affiliations:** Department of Epidemiology, Emory University; Department of Research and Public Health, Reims Teaching Hospitals, Robert Debré Hospital, Reims, France; Bridge HIV, San Francisco Department of Public Health; Department of Medicine, University of California San Francisco; HIV Epidemiology Section, San Francisco Department of Public Health

## Abstract

**Background:** Key components of *Ending the HIV Epidemic* (EHE) plan include increasing HIV antiretroviral therapy (ART) and HIV preexposure prophylaxis (PrEP) coverage. One complication to addressing this service delivery challenge is the wide heterogeneity of HIV burden and healthcare access across the U.S. It is unclear how the effectiveness and efficiency of expanded PrEP will depend on different baseline ART coverage.

**Methods:** We used a network-based model of HIV transmission for men who have sex with men (MSM) in San Francisco. Model scenarios increased varying levels of PrEP coverage relative under current empirical levels of baseline ART coverage and two counterfactual levels. We assessed the effectiveness of PrEP with the cumulative percent of infections averted (PIA) over the next decade and efficiency with the number needed to treat (NNT) by PrEP required to avert one HIV infection.

**Results:** In our projections, only the highest levels of combined PrEP and ART coverage achieved the EHE goals. Increasing PrEP coverage up to 75% showed that PrEP effectiveness was higher at higher baseline ART coverage with the PIA ranging from 61% in the lowest baseline ART coverage population to 75% in the highest ART coverage. The efficiency declined with increasing ART (NNT range from 41 to 113).

**Conclusions:** Improving both PrEP and ART coverage would have a synergistic impact on HIV prevention even in a high baseline coverage city like San Francisco. Efforts should focus on narrowing the implementation gaps to achieve higher levels of PrEP retention and ART sustained viral suppression.

## INTRODUCTION

In February 2019, the *Ending the HIV Epidemic* (EHE) plan was launched with the goal of reducing HIV incidence by 75% in 5 years and 90% in 10 years [1]. HIV antiretroviral therapy (ART) has proven highly effective at preventing transmission from an HIV-infected individual with undetectable HIV viral load [2,3]. Globally, the treatment as prevention (TasP) strategy has greatly reduced HIV incidence in high-prevalence countries [4–11]. Achieving EHE incidence reduction targets will likely require meeting high ART care cascade targets (the proportion of HIV infected individuals diagnosed, the proportion of those linked to care, and the proportion of those virally suppressed) but also expanded use of primary prevention tools like preexposure prophylaxis (PrEP) [12,13]. Men who have sex with men (MSM), accounting for two-thirds of U.S. cases in 2018 [14], are a high-priority group for PrEP expansion. It is unclear how ART and PrEP would be optimally deployed in combination to MSM to reach the EHE goals.

Addressing this implementation science question at a national level is complicated by the heterogeneity in HIV burden and healthcare access. For example, in Atlanta, knowledge of HIV status among people living with HIV (PLWH) was estimated at 84%; of the diagnosed, 81% were linked to care within one month and only 60% had suppressed viral load [14]. In contrast, in San Francisco knowledge of HIV status was estimated at 94%, with 92% of the diagnosed linked to care within one month and 77% with suppressed viral load [15]. San Francisco also had higher PrEP usage, with 45%–60% of indicated MSM using PrEP in the past year versus 15%–28% in Atlanta [16,17]. Some fear that disparities in access to and retention in PrEP will reduce its potential benefit among those who need it the most [18–20]. Understanding this local ART and PrEP epidemiological context could help policymakers select the best implementation strategy.

Mathematical modeling can support implementation decisions by projecting the population impact of different intervention scale-up strategies, but models are often limited in representing the local context.

PrEP modeling studies in the U.S. have typically evaluated settings where HIV care is limited, finding that PrEP is the most efficient when HIV incidence is high [21,22]. Fewer models have addressed the potential decrease in effectiveness or efficiency of PrEP in high ART coverage settings [23,24]. One expects a reduced PrEP effectiveness (lower proportion of infections averted) and a reduced PrEP efficiency (higher number needed to treat) in such settings. Mechanistically, this would be driven by redundant antiretroviral coverage within serodiscordant sexual partnerships, with the HIV-positive partner on ART and the HIV-negative on PrEP concurrently. Understanding this interaction would help answer whether greater priority should be to continue closing the HIV care cascade gap or to accelerate efforts to increase PrEP coverage.

In this study, we used a network-based mathematical model of MSM in San Francisco to understand the future expansion of PrEP under conditions of already relatively high ART coverage. Our primary research objective was to assess how an increase in PrEP coverage in this setting could further reduce HIV incidence based on current levels of PrEP and ART use there. Secondarily, we evaluated the general relationship between PrEP effectiveness and efficiency under different baseline ART coverage to infer the mechanisms affecting PrEP impact under varied baseline HIV treatment environments.

## METHODS

### Study Design

Our network-based mathematical model of HIV transmission dynamics was built with the EpiModel platform [25], which simulates epidemics over dynamic contact networks under the statistical framework of temporal exponential random graph models (TERGMs) [26]. We extended our previous applications estimating the impact of integrated HIV prevention and care continuum for MSM in Atlanta [22,27–29]. The current model was calibrated to the San Francisco MSM population. Full methodological details are provided in **Supplemental Technical Appendix**.

### HIV Transmission and Progression

This model simulates the dynamics of main, casual, and one-time sexual partnership for black, Hispanic, and white/other MSM, aged 15 to 65, in San Francisco. The starting network size in model simulations was 10,000 MSM, which could stochastically increase or decrease over time based on arrival (sexual debut) and departure (mortality or assumed sexual cessation at age 65).

We used primary data from the ARTnet study to fit statistical models for the TERGMs. ARTnet was a web-based egocentric network study conducted in 2017–2019 of MSM in the US, with data from 4,904 respondents reporting on 16,198 partnerships [30]. We included a main effect for geography of residence (city of San Francisco versus all other areas) in models to represent our study target population.

Parameters were weighted by census-based race/ethnicity and age distributions to account for sampling biases. In the TERGMs, predictors of partnership formation included partnership type, degree distributions by partnership types, heterogeneity in degree and assortative mixing by race/ethnicity, and age and mixing by sexual position. Partnership durations were modeled for main and causal partnerships as a set of dissolution rates stratified by partnership type and age group. Other statistical models were fit to ARTnet data to predict frequency of acts within partnerships and the probability of condom use as a function of race/ethnicity, age, diagnosed HIV status, and partnership type. MSM progressed through HIV disease with HIV viral loads modeled as a continuous attribute. Men could be screened for HIV and initiate ART, which would lower their HIV viral load (VL) and increase their longevity [31,32]. Lower VL with sustained ART use was associated with a reduced rate of HIV transmission probability per act [33,34]. Factors modifying the HIV acquisition probability per act included PrEP use [35], condom use [36], sexual position [37], and circumcision [38].

### Baseline ART and PrEP Cascades

The calibration of model parameters was done by matching model outputs to 2018 data from San Francisco (**Supplemental Tables 10–15**). HIV screening rates as well as ART starting, stopping, and restarting rates were calibrated to match surveillance statistics stratified by race: proportion of infected diagnosed, proportion of diagnosed linked to care within 3 months, proportion of treated with viral load (VL) < 200 copies/mL, and proportion of treated with VL < 200 copies/mL for at least a year (**Supplemental Table 12**).

The HIV prevention continuum consisted of initiation, adherence, and retention in PrEP care for daily oral TDF/FTC [22]. HIV-negative MSM were eligible to start if they had screened negative for HIV that week and met indications for PrEP based on CDC guidelines [39]. After PrEP initiation, differential adherence was modeled, with 78.4% reaching a high-adherence level [40] that resulted in a 99% relative reduction in HIV acquisition risk [35]. MSM with high PrEP adherence reduced their condom use, based on ARTnet estimates (see **Appendix Section 4.2**). In addition to spontaneous discontinuation, MSM stopped PrEP if they no longer exhibited PrEP indications [39]. PrEP starting and stopping rates were calibrated to match the proportion of MSM that used PrEP for any amount of time in the past 12 months (45%) and proportion of MSM who were retained on PrEP at 6 months (51%) according to 2017 San Francisco data [17] (**Supplemental Table 13**). The resulting calibrated PrEP coverage, defined as the point prevalence of PrEP users among PrEP indicated at any time point, was 25%.

### Intervention Scenarios

Our primary set of scenarios consisted of increasing PrEP coverage in San Francisco, from 25% (calibrated estimate) to 37.5%, 50%, 62.5%, and 75% (**Table 1**). We simulated these coverage levels by modifying only the spontaneous PrEP discontinuation probability, leaving all other parameters fixed. The 75% coverage was achieved by setting the spontaneous discontinuation probability to zero. However, a PrEP user could still discontinue if indications lapsed, or HIV infection or mortality occurred. This assumed a PrEP intervention targeting PrEP retention [41], based on the already high PrEP initiation rate among San Francisco MSM [17]. The reference scenario was the one calibrated on San Francisco population, with 25% PrEP coverage (**Table 1**).

**Table 1.**
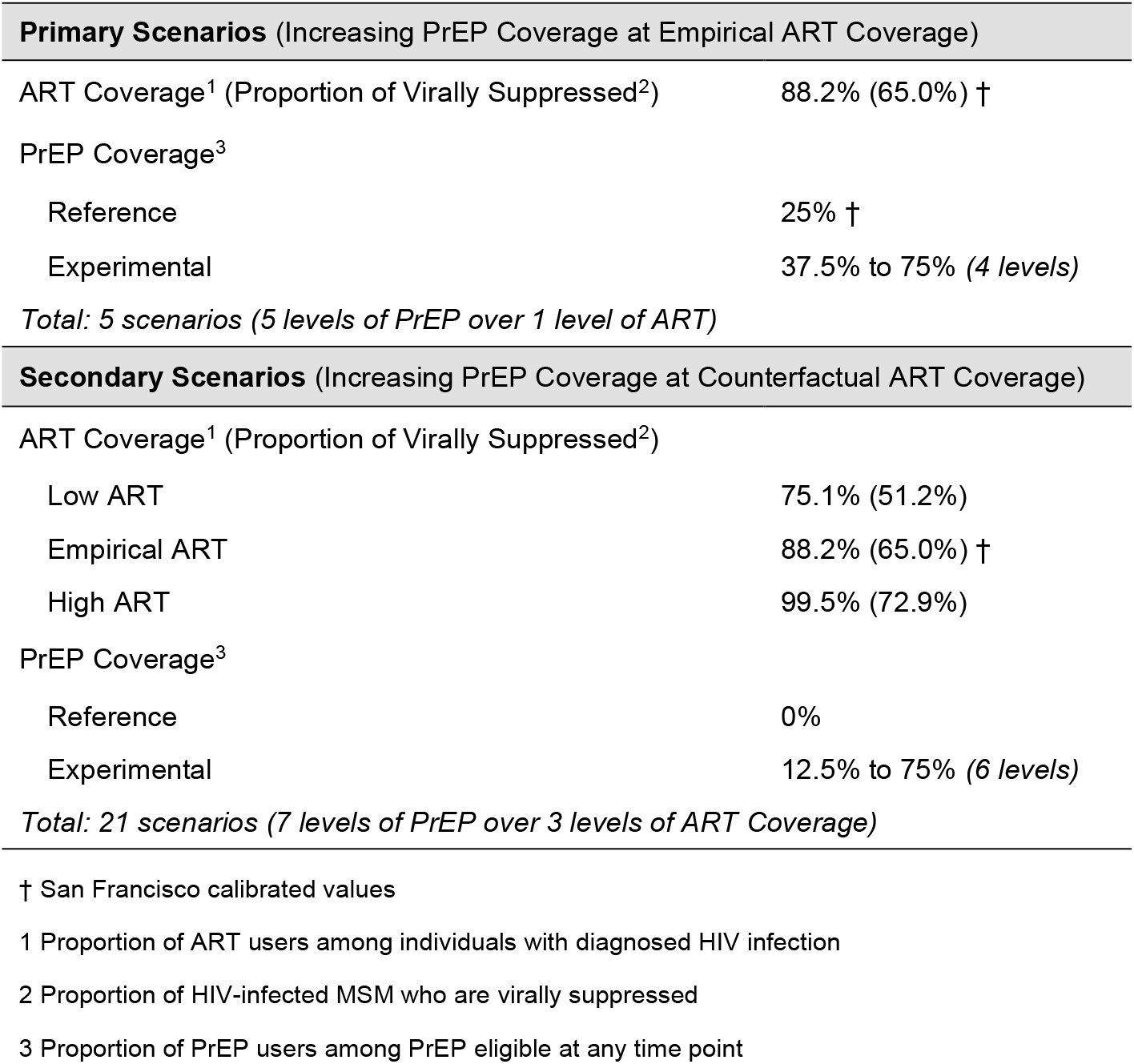
Description of the Model Scenarios

| **Primary Scenarios** (Increasing PrEP Coverage at Empirical ART Coverage) |  |
| --- | --- |
| ART Coverage<sup>1</sup> (Proportion of Virally Suppressed<sup>2</sup>) | 88.2% (65.0%) † |
| PrEP Coverage<sup>3</sup> |  |
| Reference | 25% † |
| Experimental | 37.5% to 75% (4 levels) |
| *Total: 5 scenarios (5 levels of PrEP over 1 level of ART)* |  |
| **Secondary Scenarios** (Increasing PrEP Coverage at Counterfactual ART Coverage) |  |
| ART Coverage<sup>1</sup> (Proportion of Virally Suppressed<sup>2</sup>) |  |
| Low ART | 75.1% (51.2%) |
| Empirical ART | 88.2% (65.0%) † |
| High ART | 99.5% (72.9%) |
| PrEP Coverage<sup>3</sup> |  |
| Reference | 0% |
| Experimental | 12.5% to 75% (6 levels) |
| *Total: 21 scenarios (7 levels of PrEP over 3 levels of ART Coverage)* |  |

† San Francisco calibrated values
2 Proportion of HIV-infected MSM who are virally suppressed
3 Proportion of PrEP users among PrEP eligible at any time point

To understand the broader impact of ART on PrEP effectiveness, we ran a secondary set of scenarios starting in populations where there was initially no PrEP and with different levels of ART coverage, defined as the proportion of ART users among individuals with diagnosed HIV infection. We used the proportion of HIV-infected MSM who were virally suppressed as outcome to define our counterfactual population, since this provided direct information on the proportion of infected MSM who remained infectious. Similar to PrEP coverage, we modeled different levels of viral suppression by modifying only the ART stopping probability, again justified by the already high rate of ART care linkage among San Francisco MSM [17]. The *Low ART* scenario had 50% of all HIV-infected MSM as virally suppressed, the *Empirical ART* scenario had 60%, and the *High ART* scenario had 68%. This translates to ART coverage of 75.1%, 88.2% and 99.5% respectively. The *High ART* scenario corresponded to a situation where stopping ART did not occur. The *Empirical ART* scenario was the calibrated value for San Francisco. Using these three starting populations, we ran the models for ten years with different PrEP parameters to get to 0%–75%, PrEP coverage after 4 years. Different PrEP scenarios were compared at each fixed ART coverage, with the 0% PrEP coverage of each ART coverage as the reference scenario (**Table 1**).

### Simulation and Analysis

For each scenario, we simulated the model 500 times and summarized the distribution of results with medians and 95% simulation intervals (95% SI). Epidemiological outcomes were HIV incidence per 100 person-years at risk (PYAR) and prevalence during the final intervention year; cumulative number and percent of infections averted (NIA and PIA) comparing the cumulative incidence over the 10 years intervention period for each scenario to that in the base scenarios; and number needed to treat (NNT) the number of additional MSM person-years on PrEP required to avert one new HIV infection. We also assessed whether each scenario was able to reach the two EHE goals of 75% incidence reduction in 5 years and 90% in 10 years [1].

## RESULTS

**Table 2** presents the outcomes of the primary intervention scenarios. Increasing the length of time on PrEP from a median of 5.7 months in the reference scenario to 10 months (generating 37.5% PrEP coverage) led to 19.3% (95% SI: 5.4%, 30.5%) infections averted over 10 years. Overall impact was highest at 60.1% (95% SI: 53.0%, 66.9%) of infections averted in the 75% coverage scenario. PrEP efficiency was negatively associated with PrEP coverage. With the NNT ranging from 58 (95% SI: 37, 188) to 71 (95% SI: 65, 81). Each additional 12.5% in PrEP coverage reduced the prevalence by approximately 1.5%, from 22.2% (95% SI: 20.7%, 24.2%) in the reference scenario to 16.9% (95% SI: 16.0%, 17.8%) in the 75% coverage scenario.

**Table 2.**
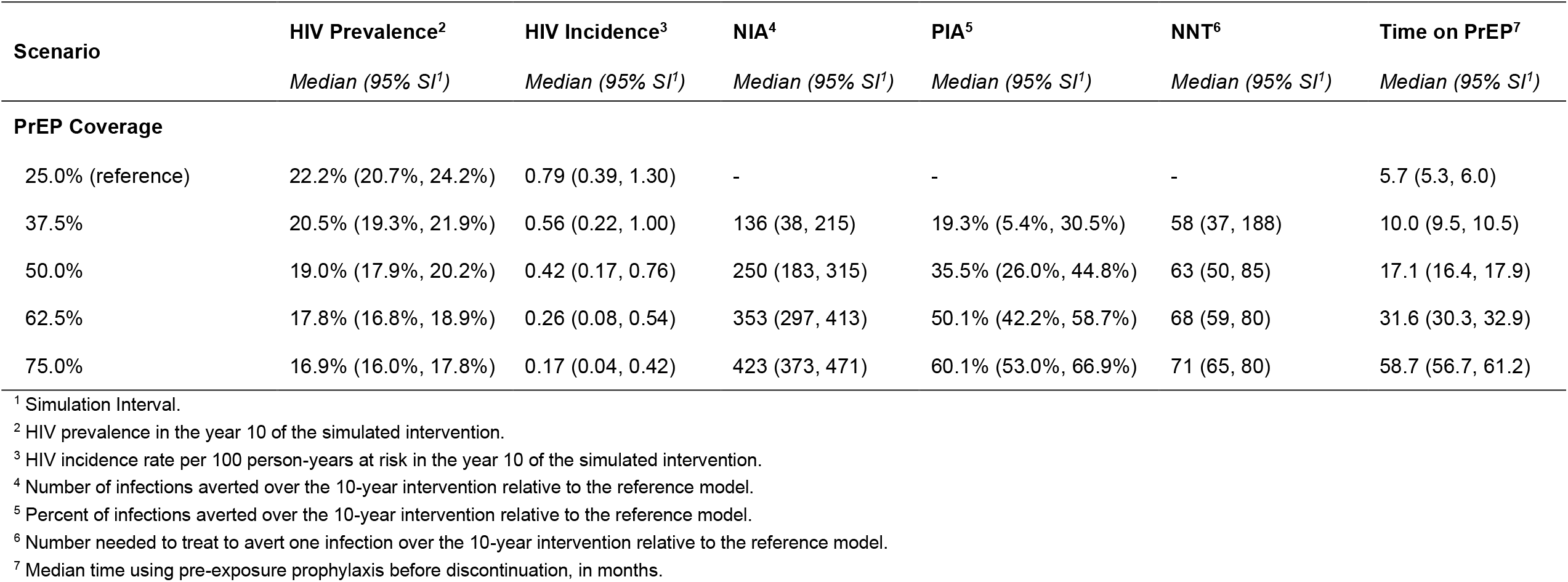
HIV Outcomes Associated with Increase to PrEP Coverage Relative to 2017 Calibrated Scenario

**Table 3** shows incidence relative to the EHE goals, with the reduction of the HIV incidence per 100 PYAR after 5 and 10 years compared to the reference scenario. None of the scenarios were able to achieve the 75% or 90% reduction at 5 and 10 years respectively through PrEP retention only and with ART coverage fixed.

**Table 3.** Reduction of HIV Incidence per 100 PYAR After 5 and 10 Years of Intervention Across Primary Scenarios

|  | **HIV Incidence Reduction<sup>1</sup>** |  |
| --- | --- | --- |
|  | *Median (95% SI<sup>2</sup>), Year 5* | *Median (95% SI<sup>2</sup>), Year 10* |
| **PrEP target** |  |  |
| 25.0% (reference) | -6% (-78%, 44%) | 0% (-67%, 50%) |
| 37.5% | 17% (-44%, 61%) | 28% (-28%, 72%) |
| 50.0% | 39% (-17%, 72%) | 44% (0%, 78%) |
| 62.5% | 56% (17%, 83%) | 67% (28%, 89%) |
| 75.0% | 67% (39%, 89%) | 78% (44%, 94%) |

<sup>1</sup> Reduction of HIV incidence rate per 100 person-years at risk compared to the first year of the simulated intervention in the no PrEP group. <sup>2</sup> Simulation Interval.

Figure 1 shows that for all levels of baseline ART coverage HIV incidence continues to decrease as PrEP coverages reaches its maximum (75%) (**Supplemental Table 20**). The 5-year EHE goal of 75% HIV incidence reduction was only achieved in the *Empirical ART* scenario for 75% PrEP coverage and in the *High ART* scenario for 62.5% and 75% PrEP coverage (**Supplemental Table 19**). The maximum incidence reduction at year 10 was 87% (95% SI: 76%, 96%) in the *High ART* scenario with 75% PrEP coverage. In the *High ART* scenario, the HIV incidence went below the epidemic control threshold (incidence < 0.1 per 100 PYAR) with PrEP coverage at 62.5% or 75%, with 0.08 (95% SI: 0.00, 0.23) and 0.04 (95% SI: 0.00, 0.15) infections per 100 PYAR at year 10 respectively (**Supplemental Table 20**).

**Figure 1.**
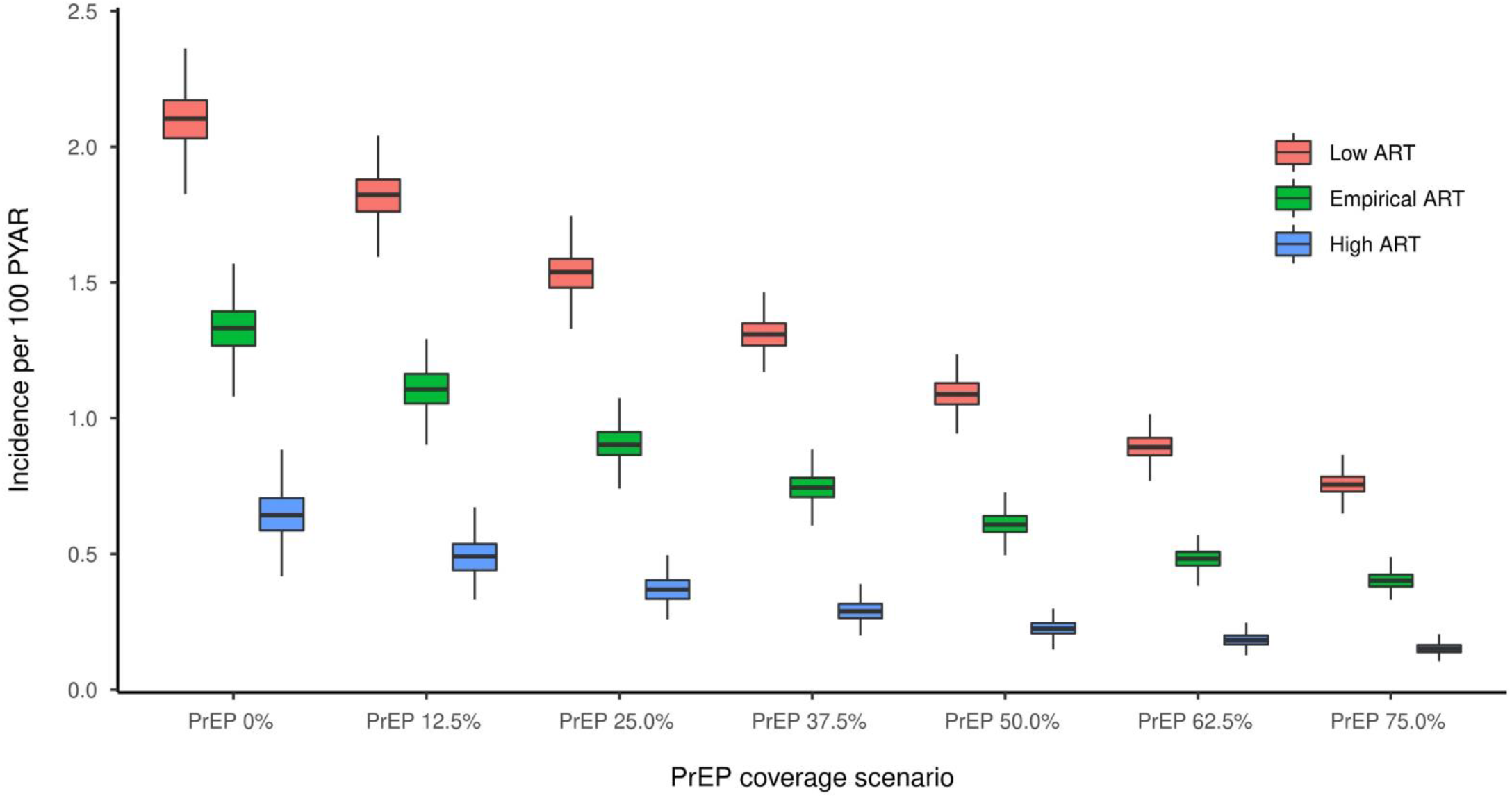
Incidence per 100 PYAR during the 10th Year of simulation for the secondary scenarios. Each color represents a baseline ART coverage, and the x axis shows the different levels of PrEP coverage, from 0% to 75%.

**Table 4** presents the NNT by PrEP to avert 1 HIV infection (NNT) as well as the absolute and relative changes in cumulative incidence by PrEP coverage in the 3 different ART coverage scenarios. As we hypothesized, PrEP efficiency was lower where ART coverage was high. At 75% PrEP coverage, the NNT in the *High ART* scenario was 113 (95% SI: 106, 124), versus 41 (95% SI: 38, 43) in the *Low ART* scenario. Similarly, the absolute number of infections averted (NIA) was lower where ART coverage was high. At 75% PrEP coverage, the NIA in the *Low ART* scenario was 892 (95% SI: 834, 953), versus 455 (95% SI: 413, 488) in the *High ART* scenarios. In contrast, with the percent of infection averted (PIA), which is standardized to the reference scenarios (0% PrEP at each baseline ART coverage), PrEP scale-up yielded better outcomes where ART coverage was high. The PIA at the highest PrEP coverage was 61% (95% SI: 57%, 66%) in the *Low ART* scenario and 75% (95% SI: 68%, 81%) in the *High ART* scenario. In these *High ART* scenarios, HIV incidence was already lower due to the prevention effect of ART and the scaling up of PrEP contributed to a larger reduction of that residual incidence.

**Table 4.** HIV Outcomes Associated with Increasing Levels of PrEP Coverage, Stratified by the Proportion of Virally Suppressed Among the Infected. For Each ART Baseline (Low, Empirical, High) PIA and NNT Are Calculated Using the no PrEP Scenario of the Baseline

| Scenario | Low ART<br>(51.2 % Viral Suppression) |  |  | Empirical ART<br>(60% Viral Suppression) |  |  | High ART<br>(72.9% Viral Suppression) |  |  |
| --- | --- | --- | --- | --- | --- | --- | --- | --- | --- |
|  | NIA<sup>2</sup> | PIA<sup>3</sup> | NNT<sup>4</sup> | NIA<sup>2</sup> | PIA<sup>3</sup> | NNT<sup>4</sup> | NIA<sup>2</sup> | PIA<sup>3</sup> | NNT<sup>4</sup> |
|  | Median<br>(95% SI<sup>1</sup>) | Median<br>(95% SI<sup>1</sup>) | Median<br>(95% SI<sup>1</sup>) | Median<br>(95% SI<sup>1</sup>) | Median<br>(95% SI<sup>1</sup>) | Median<br>(95% SI<sup>1</sup>) | Median<br>(95% SI<sup>1</sup>) | Median<br>(95% SI<sup>1</sup>) | Median<br>(95% SI<sup>1</sup>) |
| **PrEP Target** |  |  |  |  |  |  |  |  |  |
| No PrEP (reference) | - | - | - | - | - | - | - | - | - |
| 12.5% | 178<br>(67, 294) | 12%<br>(5%, 20%) | 36<br>(22, 90) | 169<br>(46, 282) | 16%<br>(4%, 27%) | 45<br>(26, 136) | 141<br>(9, 252) | 23%<br>(1%, 42%) | 64<br>(35, 435) |
| 25.0% (SFO Level) | 362<br>(264, 477) | 25%<br>(18%, 33%) | 36<br>(27, 49) | 326<br>(217, 419) | 31%<br>(20%, 39%) | 48<br>(37, 71) | 251<br>(154, 331) | 42%<br>(25%, 55%) | 75<br>(57, 120) |
| 37.5% | 516<br>(425, 594) | 35%<br>(29%, 41%) | 37<br>(33, 44) | 449<br>(370, 529) | 42%<br>(35%, 50%) | 51<br>(43, 61) | 326<br>(250, 392) | 54%<br>(41%, 65%) | 85<br>(71, 109) |
| 50.0% | 664<br>(580, 738) | 46%<br>(40%, 51%) | 38<br>(35, 43) | 557<br>(490, 628) | 52%<br>(46%, 59%) | 54<br>(48, 61) | 386<br>(321, 437) | 64%<br>(53%, 72%) | 94<br>(83, 113) |
| 62.5% | 798<br>(729, 860) | 55%<br>(50%, 59%) | 40<br>(37, 43) | 656<br>(596, 712) | 62%<br>(56%, 67%) | 57<br>(53, 62) | 425<br>(381, 469) | 70%<br>(63%, 78%) | 106<br>(96, 117) |
| 75.0% | 892<br>(834, 953) | 61%<br>(57%, 66%) | 41<br>(38, 43) | 720<br>(674, 765) | 68%<br>(64%, 72%) | 60<br>(56, 64) | 455<br>(413, 488) | 75%<br>(68%, 81%) | 113<br>(106, 124) |

<sup>1</sup> Simulation Interval.
<sup>2</sup> Number of infections averted over the 10-year intervention relative to the reference model.
<sup>3</sup> Percent of infections averted over the 10-year intervention relative to the reference model.
<sup>4</sup> Number needed to treat to avert one infection over the 10-year intervention relative to the reference model.

This increased PIA at higher ART coverage was explained by the two mechanisms by which PrEP reduces the incidence. In addition to the direct effect of preventing infections during otherwise infectious sexual contacts, PrEP has a retroactive effect through the reduction of the number of infectious MSM in the population. Infectious MSM are either newly infected MSM not yet on ART or previously virally suppressed MSM that became infectious again by dropping out of ART. In our models, we varied ART coverage only by modifying ART retention, with higher ART coverage implying a larger proportion of infectious MSM being newly infected MSM not yet on ART. Finally, as PrEP can only prevent new infections and not ART drop out, implementing it in a setting where most infectious MSM are newly infected and not yet on ART gives it the greatest opportunity to reduce the size of this population of infectious MSM. **Appendix Section 11** presents a detailed illustration of this mechanism.

## DISCUSSION

In this study, we used mathematical modeling to project the expansion of PrEP under conditions of already relatively high ART coverage among MSM in San Francisco. Given this current ART coverage level, increasing PrEP coverage from 25%, the current estimates, to 75%, would continue to significantly decrease HIV incidence, but not enough to reach Ending the HIV Epidemic (EHE) goals [1]. In the secondary scenarios where we varied the baseline ART coverage, we observed a decrease in PrEP efficiency (higher NNT) when the baseline ART coverage was higher. On the other hand, and contrary to our initial hypothesis, PrEP yielded a greater *relative* benefit (higher PIA) at higher baseline ART coverage. Our findings suggest that closing the HIV care cascade gap and accelerating efforts at PrEP coverage should be viewed as synergistic tools instead of competing options.

To reach higher PrEP coverage in a setting like San Francisco, where PrEP use is already relatively high [42], improving both uptake among priority populations and overall PrEP retention is required.

Although a challenge in higher-risk populations [43–45], several interventions have been proven effective at improving the PrEP cascade [46–48]. Our model suggests that in an epidemiological context like San Francisco, where HIV incidence has declined since the 1990s and HIV prevalence has fallen since 2012 [42], increasing PrEP retention would still further reduce HIV incidence. We show that even if PrEP efficiency would decline as ART coverage increased, it yields a greater *relative* benefit (as quantified by the PIA) when the baseline incidence is lower. The synergistic effects of PrEP and ART suggest that these complementary biomedical HIV prevention strategies should continue to be promoted simultaneously to improve population health. However, our model suggest that this may not be sufficient to reach the EHE goals unless very high PrEP and ART coverage levels are attained.

Other models have explored the overlap of PrEP in conjunction with other prevention tools. LeVasseur [23] demonstrated that the impact of PrEP would be reduced if used in conjunction with various interventions. Because their model compared several prevention strategies to no prevention strategy, it is challenging to extrapolate what effect the expansion of PrEP could be in settings with existing interventions in place. Singleton [24] demonstrated in models of Atlanta MSM that with high ART coverage, PrEP expansion would still meaningfully reduce HIV incidence with only minimal changes to efficiency. The limited changes in efficiency likely stems from their potentially unrealistic assumption of keeping PrEP retention fixed between all scenarios and systematically replacing each model agent who discontinued PrEP with another eligible agent to keep the PrEP coverage deterministically fixed. Our San Francisco model similarly projected a large percentage of infections averted when increasing PrEP at current levels of ART coverage. We further found that at higher levels of ART coverage in the population, scaling up PrEP could reduce a larger portion of the residual incidence.

Finally, we also demonstrated that the absolute number of infections averted was lower at higher levels of ART coverage; however, this metric does not consider the lower baseline incidence rate achieved by the improvement in ART coverage. For example, reducing incidence from 0.5 per PYAR to 0.1 (an absolute difference of -0.4 and a relative reduction of 83%) impacts the two metrics in an opposite way from reducing incidence from 2.3 to 0.6 (an absolute difference of -1.7 and a relative reduction of 74%). Thus, the absolute number of infections averted fails to capture how PrEP affects the residual incidence where ART coverage is already high. Because we modified ART coverage through improvements to ART retention only, our observations of increased effectiveness were a joint function of a change in the composition of infectious MSM. By increasing ART retention, most infectious MSM are newly infected MSM not yet on ART, and only a minority are MSM returning to an infectious status by dropping out of ART. Because PrEP can only prevent new infections and not ART drop out, its *relative* effectiveness grows when the proportion of newly infected MSM among infectious MSM grows. Finally, we assessed that PrEP efficiency would drop at higher ART coverage because the incidence was already lower; this generates fewer infections to be prevented by PrEP and an eligible population of roughly the same size. In summary, in settings with a high ART coverage, PrEP prevented a larger *proportion* of the HIV infections but at the cost of a larger number needed to treat by PrEP to avert one HIV infection.

## Limitations

Our models considered PrEP and ART coverage changes only through the modification of retention in those respective clinical services. This decision was based on the observation that the initiation rates for both treatments were already very high among San Francisco MSM [17]. Additionally, although our model is structured and calibrated to the unique racial/ethnic and age structure of our target population, we did not consider disparities in PrEP access among our population. Our results therefore might be overly optimistic, as the population most in need of PrEP often has the least access to it [18–20]. However, modelling such disparities would increase the complexity of our methods and the amount of data required. Finally, our study only focused on the second and third pillar of the EHE plan, ART and PrEP. This allowed us to better understand the relationship between these two prevention strategies, but meeting the EHE goals will require addressing other EHE pillars focused on improvements to HIV screening and HIV outbreak response [1].

## Conclusions

Our findings reaffirm that PrEP continues to be a critical strategy for the prevention of acquisition of HIV infection even in settings where ART coverage is already high because these two interventions can have a synergistic impact. Our study also supports the idea that ART and PrEP on their own may not be enough to reach the ambitious goals of the EHE plan without substantial scale-up of both. Given the disruptions in HIV clinical and prevention services due to the COVID-19 pandemic impact [49], it is of utmost importance to continue to tackle the HIV epidemic with all available tools even in lower incidence, higher coverage areas like San Francisco.

## Data Availability

Model code and data are available at the Github repositories linked in the Supplemental Appendix document.

https://github.com/EpiModel/SFO_PrEP

