## Supplemental Appendix for "Population Impact and Efficiency of Improvements to HIV PrEP Under Conditions of High ART Coverage among San Francisco Men Who Have Sex with Men"

Adrien Le Guillou,MD[1,2] Susan Buchbinder,MD[3] Hyman Scott,MD[3]
Albert Liu,MD[3] Diane Havlir,MD[4] Susan Scheer,PhD[3] Samuel M. Jenness,PhD[1]

[1] Department of Epidemiology, Emory University
[2] Department of Research and Public Health, Reims Teaching Hospitals, Robert Debré Hospital, Reims, France
[3] Bridge HIV, San Francisco Department of Public Health
[4] Division of Infectious Diseases, Department of Medicine, University of California San Francisco

### 1 INTRODUCTION

This supplementary technical appendix describes the mathematical model structure, parameterization, and statistical analysis of the accompanying paper in further detail.

#### 1.1 Model Framework

The mathematical models for HIV transmission dynamics presented in this study are network-based transmission models in which uniquely identifiable sexual partnership dyads were simulated and tracked over time. This partnership structure is represented using temporal exponential-family random graph models (TERGMs), described in Section 3. On top of this dynamic network simulation, the epidemic model represents demography (entries, exits, and aging), interhost epidemiology (disease transmission), intrahost epidemiology (disease progression), and clinical epidemiology (disease diagnosis and treatment and prevention interventions). Individual attributes related to these processes are stored and updated in discrete time over the course of each epidemic simulation.

The modeling methods presented here utilize and extend the *EpiModel* software platform to incorporate HIV-specific epidemiology and transmission dynamics. The HIV extensions for men who have sex with men (MSM) were originally developed by Goodreau et al. for use in prior modeling studies of MSM in the United States and South America,[1–3] and subsequently used to model for HIV preexposure prophylaxis (PrEP) among US MSM.[4–7] The most recent innovation in our modeling platform has been to incorporate primary data from the ARTnet study of MSM in the United States directly into the workflow for parameterizing the network and behavioral components.[8]

#### 1.2 Model Software

The models in this study were programmed in the R and C++ software languages using the *EpiModel* [http://epimodel.org/] software platform for epidemic modeling. *EpiModel* was developed by the authors for simulating complex network-based mathematical models of infectious diseases, with a primary focus on HIV and sexually transmitted infections (STIs).[9] *EpiModel* depends on *Statnet* [http://statnet.org/], a suite of software in R for the representation, visualization, and statistical analysis of complex network data.[10]

*EpiModel* allows for a modular expansion of its built-in modeling tools to address novel research questions. We have developed a set of extension modules into a software package called *EpiModelHIV*. This software is available for download, along with the scripts used in the execution of these models. The tools and scripts to run these models are contained in two GitHub repositories:

- [http://github.com/statnet/EpiModelHIV] contains the general extension software package. Installing this using the instructions listed at the repository homepage will also load in *EpiModel* and the other dependencies. We use a branching repository architecture on Github; the branch of the repository associated with this research project is *SFO_PrEP.*
- [http://github.com/EpiModel/SFO_PrEP] contains the scripts to execute the models and to run the statistical analyses provided in the manuscript.

##### 1.3 Core Model Specifications

We started with a network size of 10,000 MSM aged 15 to 65 to represent the larger population of sexually active MSM in San Francisco. The population size was allowed to increase and decrease with arrivals into the sexually active population at age 15 and departures related to mortality or aging out of the sexually active population at age 65. MSM were stratified by black, Hispanic, and white/other (hereafter in the text, called white) race/ethnicity in proportions equivalent to Census-derived proportions. Further details on the demography (race and age) are provided in Section 5. We used a three-stage simulation framework, first calibrating the model to diagnosed HIV prevalence and HIV care continuum parameters for 65 years of burn-in time (Stage 1), then calibrating the model to current estimated levels of PrEP coverage for 4 years of burn-in time (Stage 2), and then simulating the reference and counterfactual intervention scenarios for 10 years (Stage 3). The time unit used throughout the simulations was one week. Unless otherwise noted, all rate-based parameters listed below are to be interpreted as the rate per week and all duration-based estimates are to be interpreted as the duration in weeks.

#### 2 THE ARTnet STUDY

This model featured an innovative parameterization design in which primary individual-level and partnership-level data were used to fit statistical models for summary statistics that were then entered into the epidemic model. The primary data source for network structure and behavioral data was the ARTnet study, described below. Wherever possible, we used primary data from this study for model parameterization, and only relied on the secondary published literature for model parameters that could be generalized across target populations (e.g., HIV natural history or clinical response parameters).

##### 2.1 Study Design

This analysis used data collected in the ARTnet study of MSM in the United States in 2017–2019.[8] MSM were recruited directly after participating in the American Men's Internet Study (AMIS),[11] a parent web-based study about MSM sexual health that recruited through banner ads placed on websites or social network applications. At the completion of AMIS, MSM were asked to participate in ARTnet, which focused on sexual network features. ARTnet data collection occurred in two waves (following AMIS): July 2017 to February 2018 and September 2018 to January 2019.

Eligibility criteria for ARTnet were male sex at birth, current male cisgender identity, lifetime history of sexual activity with another man, and age between 15 and 65. Respondents were deduplicated within and across survey waves (based on IP and email addresses), resulting in a final sample of 4904 participants who reported on 16198 sexual partnerships. The Emory University Institutional Review Board approved the study.

##### 2.2 Primary Measures

ARTnet participants were first asked about demographic and health-related information. Covariates used in this analysis included race, age, ZIP Code of residence, and current HIV status. ZIP Codes were transformed into Census regions/divisions and urbanicity levels by matching against county databases (using standardized methods for selecting county in the small number of cases when ZIP Codes crossed county lines). Participants reporting as never testing for HIV, having indeterminate test results, or never receiving test results were classified as having an unknown HIV status.

Participants were then asked detailed partner-specific questions for up to their most recent 5 partners. The detailed partner-specific questions included attributes of the partner and details about the partnership itself. Partner attributes considered here included age, race/ethnicity, and HIV status. Participants were allowed to report any partner attribute as unknown. When partner age was unknown, age was imputed based on a response to a categorical question (e.g., 5–10 years younger/older, 2–5 years younger/older). Partnerships were classified into three types: main (respondent reported they considered this partner a “boyfriend, significant other, or life partner”) casual (someone they have had sex with more than once, but not a main partner), and one-time.[12] For one-time partners, we asked for the date that sexual activity occurred. For persistent (main and casual) partnerships, we asked for the date of most recent sex, the date first sex (which could have been prior to the past year), and whether the partnership was ongoing (if the participant expected sexual activity would occur in the future). For each partnership, we asked whether (for one-time) or how frequently (for persistent) anal sex occurred.

Outcome measures include descriptive statistics for characteristics of participants and their reported partnerships, and the aggregate network statistics used to fit the TERGMs underlying epidemic simulations on dynamic networks. The network statistics include ego degree, attribute mixing in partnerships, and the current length of ongoing partnerships, stratified by the attributes of persons and partnerships. Degree is a property of individuals, whereas mixing and length are properties of partnerships. Degree was defined as the ongoing number of persistent partners measured on the day of the survey (includes main and casual partnerships). Degree is not defined for one-time partnerships, so for these we instead calculated a weekly rate of new contacts by subtracting the total main and casual partners from the total past-year partners, and dividing by 52. Partnership length for ongoing main and casual partnerships was calculated by taking the difference between the survey date and the partnership start date. The mean length of ongoing partnerships is the network statistic needed for TERGM estimation; the logic and derivation are explained here.[9] Mixing was measured by the relative frequency of partnerships that occurred within and between groups defined by race/ethnicity, and age.

##### 2.3 Statistical Analysis

We fit a series of generalized linear models (GLMs) to estimate summary statistics for features of the sexual network structure and the behavior within partnerships. Specific GLM parameterizations are detailed below in the discussion of each set of model parameters. Common across all models was the general approach of including geography of residence as a main effect with two levels (Atlanta versus all other areas). This allowed for the model coefficients and predicted summary statistics to vary by geography while ensuring stability of outcomes under the assumption of conditional exchangeability.

#### 3 NETWORKS OF SEXUAL PARTNERSHIPS

We modeled networks of three interacting types of sexual relations: main partnerships, casual (but persistent) partnerships, and one-time anal intercourse contacts. We first describe the methods conceptually, including the parameters used to guide the model and their derivation, and then present the formal statistical modeling methods. Consistent with our parameter derivations, all relationships are defined as those in which anal intercourse is expected to occur at least once.

##### 3.1 Conceptual Representation of Sexual Networks

Our modeling methods aim to preserve certain features of the cross-sectional and dynamic network structure as observed in our primary data, while also allowing for mean relational durations to be targeted to those reported for different groups and relational types. Our methods do so within the context of changing population size (due to births, deaths, arrivals and departures from the population) and changing composition by attributes such as age. The broader motivation, methodological details, and link between models and primary data are described here.[9]

The network features that we aim to preserve are as follows:

- Persistent (Main and Casual) Partnerships
    - The mean degree (number of ongoing partners) stratified by main and casual partnership types, and the proportion of men with concurrency (2 or more ongoing partners) for each partnership type, at any time point.
    - Variations in the mean degree specific to each persistent partnership type by:
        - Race/ethnicity group (3 categories for black, Hispanic, and white MSM).
        - Age group (5 categories for 15–24, 25–34, 35–44, 45–54, and 55–64).
        - Cross-type degree: Degree in the other persistent partnership type (e.g., mean degree of MSM for main partnerships given current casual degree of 0, 1, 2, 3).
    - Selection of partners within the same race/ethnicity group (mixing by race/ethnicity).
    - Selection of partners within the same age group (mixing by age).
    - Mean partnership durations, stratified by main and casual partnership types, and by mixing within age groups.
- One-Time Partnerships
    - The overall rate of having one-time anal intercourse partnerships per week.
    - Variations in this contact rate by:
        - Race/ethnicity group.
        - Age group.
        - Total persistent degree (sum of main and casual partnerships ongoing).
        - Risk level heterogeneity and beyond the risk heterogeneity associated with demographics (mean partnership rates for five quintiles of MSM stratified by mean one-time rates).

    - Selection of partners within the same race/ethnicity group (mixing by race/ethnicity).
    - Selection of partners within the same age group (mixing by age).
- Common to Persistent and One-Time Partnership Types
    - Prohibitions against MSM with incompatible sexual positioning roles (e.g., no partnerships between exclusively receptive MSM).

##### 3.1.1 Overall Mean Degree for Persistent Partnerships

Ongoing persistent partnerships (whether main or casual) were defined from the partnership-level ARTnet dataset as those in which sex had already occurred more than once, and in which the respondent anticipated having sex again. The momentary main or casual mean degree is then defined as the mean of the degree of all MSM for main or casual partnerships on the day of study. We estimated this with a Poisson model with main or casual degree as the outcome and a dummy variable for San Francisco residence as the predictor and then exponentiating the coefficients, resulting in an estimated mean main degree of 0.472 and a mean casual degree of 0.882.

In addition, we modeled the proportion of MSM with concurrency (degree of 2 or more) by partnership type. This was estimated with logistic regression models for binary outcomes with a dummy variable for San Francisco residence as the predictor. Taking the inverse of the logit of the coefficient yielded the predicted probabilities of 2.4% for main concurrency and 26.0% for casual concurrency.

##### 3.1.2 Heterogeneity in Mean Degrees for Persistent Partnerships

We fit the heterogeneity in main and casual mean degree by fitting three Poisson regression models. For race/ethnicity, we estimated the mean degree for each group within the target population by including dummy variables for city and race/ethnicity. For age, we modeled the non-linear relationship between age and mean degrees by including city, age group, and square root of age group to allow for a non-linear relationship between age and the outcome. For cross type degree, we modeled the mean degree for main partnerships as a function of degree of casual partnerships, and vice versa, again with city also as a predictor. For each of the 6 models (2 partnership types times three predictors of interest), we estimated the statistical models and then exponentiated the coefficients to obtain the rates for each stratum. Those are shown in the **Supplemental Table 1** below.

| **Supplemental Table 1.** Heterogeneity in Mean Main and Casual Degree by Race/Ethnicity, Age Group, and Cross Type Degree of Ego (Respondent) | | |
|---|---|---|
| **Predictor** | **Main Mean Degree** | **Casual Mean Degree** |
| **Race/Ethnicity** | | |
| Black | 0.321 | 1.001 |
| Hispanic | 0.488 | 0.848 |
| White | 0.475 | 0.882 |
| **Age Group** | | |
| 15–24 | 0.452 | 0.463 |
| 25–34 | 0.567 | 0.749 |
| 35–44 | 0.542 | 0.960 |
| 45–54 | 0.450 | 1.095 |
| 55–64 | 0.341 | 1.156 |
| **Cross Type Degree** | | |
| 0 | 0.562 | 1.061 |
| 1 | 0.449 | 0.674 |
| 2 | 0.358 | 0.428 |
| 3 | 0.286 | — |

###### *3.1.3 Mixing by Race/Ethnicity and Age for Persistent Partnerships*

Respondents reported on their perception of the race and ethnicity (Hispanic/non-Hispanic) for each partner. We categorized the respondents' and partners' races into three mutually exclusive groups: black, Hispanic, and white. Using logistic regression models, we estimated the proportion of partnerships that were between MSM of the same race (within-group mixing) by evaluating relationship between the respondent group and partner group as a binary outcome (using the geography of residence predictor as a main effect with two levels, Atlanta versus all other areas). The inverse logit of the coefficients is then interpreted as the predicted probability of a same race/ethnicity partnership. The values were 76.5% for main partnerships and 63.3% for casual partnerships.

For mixing by age, we used a model parameterization for the 5-category age group that allowed for differences in the level of age mixing that could vary by age group (differential homophily). We fit a logistic regression model for partnerships, with being in a partnership of the same age group as the outcome and the age group of the respondent as the main predictor. With the inverse logit transformation, the probabilities of partnerships within the same age group, stratified by partnership type are shown in the **Supplemental Table 2** below.

| **Supplemental Table 2.** Proportion of Main and Casual Partnerships within the Same Age Group, by Age of Ego (Respondent) | | |
|---|---|---|
| **Age Group** | **Main Within Group** | **Casual Within Group** |
| 15–24 | 74.6% | 58.8% |
| 25–34 | 65.4% | 48.1% |
| 35–44 | 54.7% | 37.6% |
| 45–54 | 43.6% | 28.1% |
| 55–64 | 33.2% | 20.2% |

###### *3.1.4 Duration of Persistent Partnerships*

We model partnership dissolution as a heterogenous, geometrically distributed process with unique parameters for each relational type. The geometric distribution for relational durations implies a "memoryless process," which is a common assumption within ordinary differential equation modeling. Although this assumption implies that the rate of dissolution does not depend on the current age of the partnership, the overall exponential shape of the dissolution distribution matches reasonably well to empirical data on relational durations. The fit is improved considerably when the partnership types are stratified, as we do here, implying a mixture of geometric distributions. Once one-time contacts are removed, and longer-duration main partnerships are separated from shorter-term causal partnerships, the set of geometric distribution fits the empirical data on partnership durations well.

The fit is improved further by stratifying based on the interaction between partnership type and the age of both members within the dyad. For this analysis, we explored how relationship duration varied by multiple demographic characteristics, and unsurprisingly age was most strongly associated with duration. For this model parameterization, we specifically elected to estimate and input parameters based on matched age groups (that is, partnerships between two persons of the same age).

As detailed in previous work,[1,9] for memoryless processes, the expected age of an extant (ongoing) relationship at any moment in time is an unbiased estimator of the expected uncensored duration of relationships, given the balancing effects of right-censoring and length bias for this distribution. Raw relational ages were calculated as the difference between first sex date and the study date for each dyad the ego reported sex with more than once in the interval. To derive our estimator of relational age, we take the median of the observed distribution and then calculate the mean for the geometric distributions associated with that median. To account for estimation within the San Francisco target population, we weighted this estimator by the inverse of the relative differences in San Francisco partnerships to non-San Francisco partnerships.

The resulting expected relational ages are summarized in **Supplemental Table 3** below.

| **Supplemental Table 3.** Duration of Main and Casual Partnerships by Dyadic Age Group of Ego (Respondent) and Alter (Partner) | | |
|---|---|---|
| **Dyadic Age Group** | **Main Relational Age (Weeks)** | **Casual Relational Age (Weeks)** |
| Both 15–24 | 69.3 | 66.7 |
| Both 25–34 | 202.9 | 98.8 |
| Both 35–44 | 392.1 | 144.4 |
| Both 45–54 | 550.8 | 232.1 |
| Both 55–64 | 816.6 | 203.2 |
| Different Groups | 261.6 | 131.9 |

###### *3.1.6 Overall Mean One-Time Contact Rate*

In addition to persistent main and casual partnerships, we modeled one-time sexual contacts involving anal intercourse based on ARTnet reports on the number and variation in these types of relations. As noted above, degree is not defined for one-time contacts, so for these we instead calculated a weekly rate of new contacts by subtracting the total main and casual partners from the total past-year partners. We estimated the weekly rate by fitting a Poisson regression model with the count of one-time contacts as a function of city, exponentiating the coefficient to get the predicted count, and dividing by 52 to get the week rate. The overall mean one-time contact rate was 0.193 anal intercourse contacts per week.

###### *3.1.7 Heterogeneity in One-Time Contact Rates*

Heterogeneity in one-time contact rates was modeled with four Poisson regression models to estimate the rates as a function of race/ethnicity, age group, risk level strata, and total persistent (main plus casual) degree. Similar to the one-time rate, we fit these models with geography of residence as a main effect (which had two levels, San Francisco versus all other areas, with the former level used for predictions) and exponentiated the coefficients and then divided by 52 to get the group-specific rates. For age group, similar to the estimation of degree, we modeled this non-linearly by including age group and the square root of age group as the joint predictors (along with city). The results are shown in **Supplemental Table 4** below.

| **Supplemental Table 4.** Weekly One-Time Contact Rates by Race/Ethnicity, Age Group, Risk Level, and Total Persistent Degree of Ego (Respondent) | |
|---|---|
| **Predictor** | **Weekly Contact Rate** |
| **Race/Ethnicity** | |
| Black | 0.155 |
| Hispanic | 0.178 |
| White | 0.197 |
| **Age Group** | |
| 15–24 | 0.117 |
| 25–34 | 0.186 |
| 35–44 | 0.222 |
| 45–54 | 0.228 |
| 55–64 | 0.213 |
| **Risk Level Quintile** | |
| 1 | 0.000 |
| 2 | 0.000 |
| 3 | 0.030 |
| 4 | 0.110 |
| 5 | 0.827 |
| **Total Persistent Degree** | |
| 0 | 0.088 |
| 1 | 0.103 |
| 2 | 0.219 |
| 3+ | 0.516 |

##### *3.1.8 Mixing by Race/Ethnicity and Age for One-Time Contacts*

We used a similar approach to within-group mixing by race/ethnicity and age group for one-time contacts to the one used for persistent contacts, with one difference that we did not model differential homophily by age group to improve model stability. Therefore, the overall proportion of one-time contacts that were within the same race/ethnic group was 71.4% and the proportion of one-time contacts that were within the same age group was 31.7%.

##### *3.1.9 Mixing by Sexual Role Across All Partnership Types*

We assign men a fixed sexual role preference (exclusively insertive, exclusively receptive, versatile). The model then includes an absolute prohibition, such that two exclusively insertive men cannot partner, nor can two exclusively receptive men. We estimated the proportion of men that were in each category (insertive, receptive, and versatile) by analyzing whether men had only insertive anal intercourse, only receptive anal intercourse, or both insertive and receptive anal intercourse (respectively) in their past five anal partnerships over the past year. These proportions were stratified (restricted) by geography of residence to the city of San Francisco. The proportions were: 18.5% exclusively insertive, 27.1% exclusively receptive, and 54.4% versatile.

##### 3.2 Statistical Representation of Sexual Networks

Exponential-family random graph models (ERGMs) and their dynamic extension temporal ERGMs (TERGMs) provide a foundation for statistically principled simulation of local and global network structure given a set of target statistics from empirical data. Main and casual relationships were modeled using TERGMs,[13] since they persist for multiple time steps. One-time contacts, on the other hand, were modeled using cross-sectional ERGMs.[14] Formally, our statistical models for relational dynamics can be represented as five equations for the conditional log odds (logits) of relational formation and persistence at time *t* (for main and casual relationships) or for relational existence at time *t* (for one-time contacts):

$$logit\left(P\left(Y_{ij,t}=1\middle|\, Y_{ij,t-1}=0,\, Y_{ij,t}^{C}\right)\right) = \theta_m^{+\prime}\partial\left(g_m^{+}(y)\right) \qquad \text{Main partnership formation}$$

$$logit\left(P\left(Y_{ij,t}=1\middle|\, Y_{ij,t-1}=0,\, Y_{ij,t}^{C}\right)\right) = \theta_c^{+\prime}\partial\left(g_c^{+}(y)\right) \qquad \text{Casual partnership formation}$$

$$logit\left(P\left(Y_{ij,t}=1\middle|\, Y_{ij,t-1}=1,\, Y_{ij,t}^{C}\right)\right) = \theta_m^{-\prime}\partial\left(g_m^{-}(y)\right) \qquad \text{Main partnership persistence}$$

$$logit\left(P\left(Y_{ij,t}=1\middle|\, Y_{ij,t-1}=1,\, Y_{ij,t}^{C}\right)\right) = \theta_c^{-\prime}\partial\left(g_c^{-}(y)\right) \qquad \text{Casual partnership persistence}$$

$$logit\left(P\left(Y_{ij,t}=1\middle|\, Y_{ij,t}^{C}\right)\right) = \theta_o{}'\partial\left(g_o(y)\right) \qquad \text{One-time contact existence}$$

where:

- $Y_{ij,t}$ = the relational status of persons *i* and *j* at time *t* (1 = in relationship/contact, 0 = not).
- $Y_{ij,t}^{C}$ = the network complement of *i,j* at time *t*, i.e. all relations in the network other than *i,j.*
- $g(y)$ = vector of network statistics in each model (the empirical statistics defined in the tables above).
- $\partial\left(g(y)\right)$ = the change in $g(y)$ when $Y_{ij}$ is toggled from 0 to 1 (for formation models) or 1 to 0 (for persistence models).
- $\theta$ = vector of parameters in the model.

For $g(y)$ and $\theta$, the superscript distinguishes the formation model (+), persistence model (-) and existence models (neither). The subscript indicates the main (m), casual (c) and one-time (o) models.

The recursive dependence among the relationships renders the model impossible to evaluate using standard techniques; we use MCMC in order to obtain the maximum likelihood estimates for the $\boldsymbol{\theta}$ vectors given the $\boldsymbol{g}(\boldsymbol{y})$ vectors.

Our method of converting the statistics laid out in Section 3.1 into our fully specified network models consists of the following steps:

1. Construct a cross-sectional network of 10,000 men with no relationships.
2. Assign men demographics (race/ethnicity and age) based on Census data for San Francisco and assign men sexual roles based on frequencies listed above, as well as one-time risk quintiles (20% of the men in each race per quintile).
3. Calculate the target statistics (i.e., the expected count of each statistic at any given moment in time) associated with the terms in the formation model (for the main and casual partnerships) and in the existence model (for one-time contacts).
4. Assign each node a place-holder main and casual degree (number of ongoing partnerships) that is consistent with the estimated distributions, and store these numbers as a nodal attribute. (Note: this does not actually require individuals to be paired up into the partnerships represented by those degrees).
5. For the main and casual networks, use the mean relational durations by age group combination to calculate the parameters of the persistence model, using closed-form solutions, given that the models are dyadic-independent (each relationship's persistence probability is independent of all others).
6. For the main and casual networks, estimate the coefficients for the formation model that represent the maximum likelihood estimates for the expected cross-sectional network structure.
7. For the one-time network, estimate the coefficients for the existence model that represent the maximum likelihood estimates for the expected cross-sectional network structure.

Steps 5–7 occur within the *EpiModel* software, and use the ERGM and STERGM methods therein. They are completed efficiently by the use of an approximation in Step 6.[15] During the subsequent model simulation, we use the method of Krivitsky[16] to adjust the coefficient for the edges term in each model at each time step, in order to preserve the same expected mean degree (relationships per person) over time in the face of changing network size and nodal composition. At all stages of the project, simulated partnership networks were checked to ensure that they indeed retained the expected cross-sectional structure and relational durations throughout the simulations.

### 4 BEHAVIOR WITHIN SEXUAL PARTNERSHIPS

In this study, we model three phenomena consecutively within relationships at each time step: the number of anal intercourse sex acts, condom use per sex act, and sexual role per sex act. We simulate these within all relationships regardless of HIV status (whether diagnosed or not).

#### 4.1 Anal Intercourse Acts Per Partnership

The rate of anal intercourse is applicable to persistent (main and casual) partnerships in which there are repeated anal intercourse acts between the start and end of the partnership. We use ARTnet data on the overall rate and predictors of variation in rates unique to each partnership type. For one-time contacts, we assumed that the number of anal intercourse exposures was one by definition, although there could have been multiple anal intercourse acts within an exposure due to role versatility (see Section 4.4). The modeling of act rates here is based on the expectation that changes in coital frequency depend on race/ethnicity, age, diagnosed HIV status, and partnership type.

##### *4.1.1 Measurement of Acts in ARTnet*

We measured the number of acts within each reported partnership within the ARTnet study by asking participants about the frequency of anal intercourse acts. Study participants could report on the average number of acts within the partnership over the past year by week, month, year, or total partnership duration. We then scaled this into a total weekly act rate. The final ARTnet partnership-level dataset on 16198 partnerships includes this weekly rate as the outcome and predictors at the individual and dyadic level that we used for statistical modeling as described below.

##### *4.1.2 Statistical Models of Act Rates*

With this partnership-level dataset, we then modeled the count of acts per year per partnership based on the Poisson regression formula:

$$Y_i \sim \beta_0 + \beta_1 X_1 + \beta_2 X_1^2 + \beta_3 X_2 + \beta_4 X_3 + \beta_5 X_1 X_3 + \beta_6 X_4 + \beta_7 X_4^2 + \beta_8 X_5 + \beta_9 X_6$$

where:

$Y_i$ = Log of the count of acts per year.

$X_1$ = Duration of partnership in weeks at the survey date.

$X_2$ = Racial/ethnic combination of the ego (respondent) and alter (partner), coded in 6 categories to capture within and across group mixing: black-black, black-Hispanic/white, Hispanic-black/white, Hispanic-Hispanic, white-black/Hispanic, white-white.

$X_3$ = Partnership type (0 = main or 1 = casual).

$X_4$ = The combined age of ego and alter in years.

$X_5$ = The concordant diagnosed HIV-positive status of both ego and alter compared to all other combinations of dyadic HIV status (1 = concordant positive; 0 = all other combinations of dyadic HIV status).

$X_6$ = Residence (1 = San Francisco; 0 = all other areas)

Note that we modeled the partnership duration and combined age of partners quadratically, and we modeled the interaction of partnership duration and partnership type. Terms within the prediction model were selected based on a combination of *a priori* theory and exploratory data analysis. The coefficients for the model, and their lower and upper 95% confidence intervals, are presented in **Supplemental Table 5** below. Exponentiating any linear combination of coefficients will yield the yearly rates, which may be converted to weekly rates through division.

**Supplemental Table 5.** Statistical Model of Act Rates in Main and Casual Partnerships

| Model Parameter | Estimate | Lower 95% CI | Upper 95% CI |
|---|---|---|---|
| $\beta_0$ (Intercept) | 5.038E+00 | 4.997E+00 | 5.079E+00 |
| $\beta_1$ (Duration) | -1.060E-03 | -1.097E-03 | -1.024E-03 |
| $\beta_2$ (Duration$^2$) | 4.910E-07 | 4.641E-07 | 5.176E-07 |
| $\beta_3$ (B-H/W Combo) | 5.019E-01 | 4.708E-01 | 5.330E-01 |
| $\beta_3$ (H-B/W Combo) | 1.551E-01 | 1.281E-01 | 1.823E-01 |
| $\beta_3$ (H-H Combo) | 2.096E-01 | 1.817E-01 | 2.376E-01 |
| $\beta_3$ (W-B/H Combo) | 3.981E-01 | 3.728E-01 | 4.235E-01 |
| $\beta_3$ (W-W Combo) | 1.797E-01 | 1.550E-01 | 2.047E-01 |
| $\beta_4$ (Casual Type) | -1.123E+00 | -1.131E+00 | -1.114E+00 |
| $\beta_5$ (Duration x Casual Type) | -5.555E-04 | -6.032E-04 | -5.081E-04 |
| $\beta_6$ (Combined Age) | -1.374E-02 | -1.468E-02 | -1.280E-02 |
| $\beta_7$ (Combined Age$^2$) | 6.932E-05 | 6.311E-05 | 7.552E-05 |
| $\beta_8$ (HIV+ Concordant) | 4.067E-01 | 3.904E-01 | 4.229E-01 |
| $\beta_9$ (Atlanta residence) | -1.512E-01 | -1.720E-01 | -1.306E-01 |

Abbreviations: CI, confidence interval; B-H/W, black ego with either a Hispanic or white alter; H-B/W, Hispanic ego with either a black or white alter; H-H, Hispanic ego with a Hispanic alter; W-B/H, white ego with either a black or Hispanic alter; W-W, white ego with a white alter.

##### 4.1.3 Predicted Rates in Epidemic Model

Predicted weekly rates of anal intercourse based on the combination of partnership and individual attributes is then obtained dynamically by predicting from the statistical model with inputs based on the current simulated population. *EpiModel* tracks the current age of partners, the duration of their partnership, their racial combination, and the partnership type. This set of predictors was input into a predict function in R to obtain the weekly mean rates in each stratum. The size of the potential set of strata and corresponding predicted means is therefore nearly infinite based on all the potential combinations of input values.

In Supplemental Figure 1 below, we display some example weekly rates based on a subset of model inputs. This figure shows that rates decline in partnerships with a longer duration, that they are higher in partnerships in which both partners are younger, they are lower for casual partnerships (ptype = 2) compared to main partnerships, and that they are higher in white-white partnerships compared to black-black partnerships. The act rates generally ranged from 0.5 acts per week to 2 acts per week. Other predicted rates may be obtained by exponentiating the coefficients in the table above and dividing by 52 (to convert from yearly rates to weekly rates).

**Supplemental Figure 1.** Predicted Weekly Act Rates from the Poisson Statistical Model, by Partnership Duration, Partnership Type (ptype: 1 = Main; 2 = Casual), Combined Partner Age (comb.age: 40 and 80 Years).

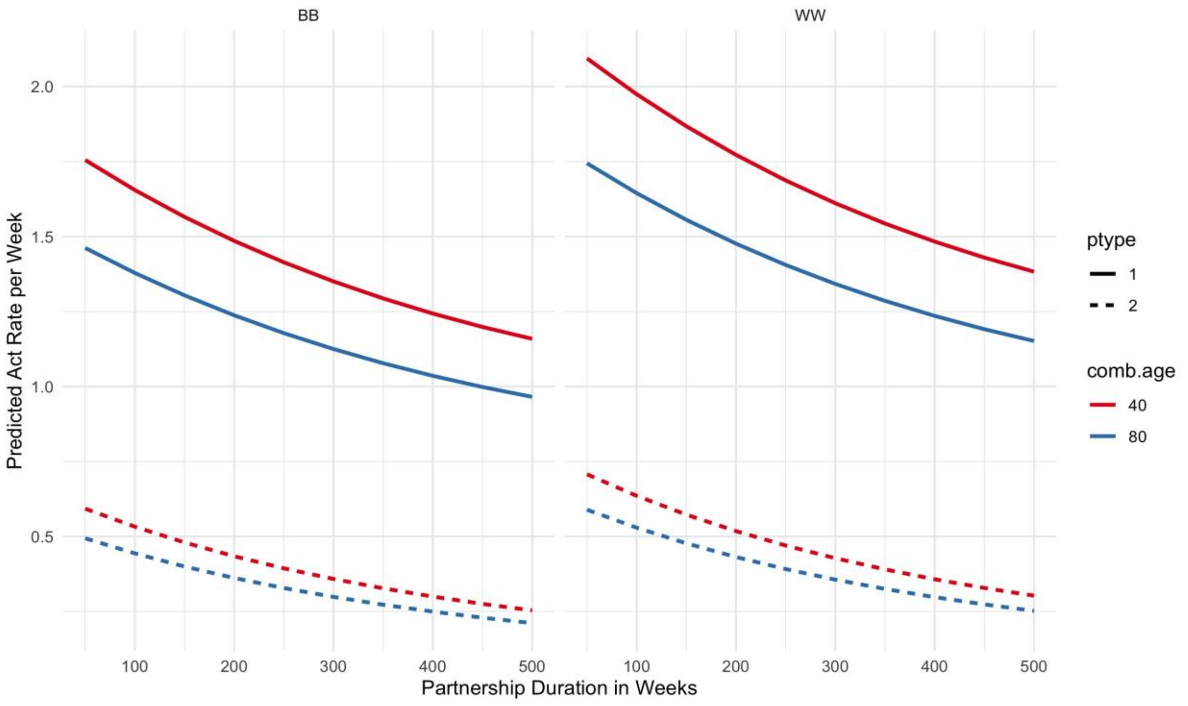

Based on these model predictions, which represent means for each linear combination, we then drew individual counts of acts per partnership per time step in *EpiModel* using the rpois function to draw randomly from the Poisson distribution with a vector of parameters, one value for each partnership.

##### *4.1.4 Cessation of Sexual Activity During Late-Stage AIDS*

In addition to these data-driven statistical calculations, we assumed that MSM in late stages of AIDS (HIV viral load above 5.75), had no acts due to active disease that would limit their sexual activity. This reflected the mid-point between set-point viral load of chronic stage infection (4.5 $\log_{10}$) and peak viral load (7.0 $\log_{10}$, corresponding to the nadir of immunological function). We had no primary data in ARTnet on sexual partnerships in this late disease stage, but prior analysis and modeling studies support a large decline in sexual activity due to AIDS.[17]

#### 4.2 Condom Use Per Act

We modeled condom use within all three partnership types (main, casual, and one-time contacts) based on ARTnet data on the frequency of condom use within reported partnerships. We followed the same general approach to measuring, fitting statistical models, and dynamically predicting condom use within *EpiModel* as we used for rates of anal intercourse. The modeling of condom use here is based on the expectation that changes in condom use depend on race/ethnicity, age, diagnosed HIV status, current PrEP use, and partnership type.

##### *4.2.1 Measurement of Condom Use in ARTnet*

We measured condom use within partnerships in the ARTnet study by asking about the frequency of condom use (for persistent partnerships) or whether condom use occurred (for one-time partnerships) during anal intercourse. Study participants first reported on the number of anal intercourse acts that occurred in the time intervals described above, and then we followed-up with a question on the number of those total acts that involved condom use. We then transformed these subsetted counts into proportions of acts that were condom-protected. This resulted in a U-shaped distribution of proportions, with most persistent partnerships involving either always or never condom use. For this current study, we simplified the outcome variable to any condom use (yes, no) over the past year.

##### *4.2.2 Statistical Models of Condom Use Probabilities*

With the outcome described above, we used the partnership-level dataset to fit two logistic regression models for any condom use in the partnership, with one model for persistent (main and casual) and another model for one-time partnerships. The linear model formula for persistent partnerships was as follows:

$$Y_i \sim \beta_0 + \beta_1 X_1 + \beta_2 X_1^2 + \beta_3 X_2 + \beta_4 X_3 + \beta_5 X_1 X_3 + \beta_6 X_4 + \beta_7 X_4^2 + \beta_8 X_5 + \beta_9 X_6 + \beta_{10} X_7$$

where:

$Y_i$ = Log odds of the probability of condom use per act.

$X_1$ = Duration of partnership in weeks at the survey date.

$X_2$ = Racial/ethnic combination of the ego (respondent) and alter (partner), coded in 6 categories to capture within and across group mixing: black-black, black-Hispanic/white, Hispanic-black/white, Hispanic-Hispanic, white-black/Hispanic, white-white.

$X_3$ = Partnership type (0 = main; 1 = casual).

$X_4$ = The combined age of ego and alter in years.

$X_5$ = The concordant diagnosed HIV-positive status of both ego and alter (1 = concordant positive; 0 = all other combinations of dyadic HIV status).

$X_6$ = Current use of pre-exposure prophylaxis (PrEP) by the ego (respondent).

$X_7$ = Residence (1 = San Francisco; 0 = all other areas).

Note that we modeled the partnership duration and combined age of partners quadratically, and we modeled the interaction of partnership duration and partnership type. Terms within the prediction model were selected based on a combination of *a priori* theory and exploratory data analysis. The coefficients for the model, and their lower and upper 95% confidence intervals, are presented in **Supplemental Table 6** below. Taking the inverse logit of the linear combination of coefficients will yield to the strata-specific predicted probabilities of condom use within the partnership.

**Supplemental Table 6.** Statistical Model of Per Act Condom Use Probability for Main and Casual Partnerships

| Model Parameter | Estimate | Lower 95% CI | Upper 95% CI |
|---|---|---|---|
| $\beta_0$ (Intercept) | 2.0226E+00 | 1.3209E+00 | 2.7241E+00 |
| $\beta_1$ (Duration) | -3.1387E-03 | -4.0168E-03 | -2.2740E-03 |
| $\beta_2$ (Duration$^2$) | 1.2494E-06 | 5.8218E-07 | 1.8543E-06 |
| $\beta_3$ (B-H/W Combo) | -3.1033E-01 | -8.2793E-01 | 2.0327E-01 |
| $\beta_3$ (H-B/W Combo) | -3.8142E-01 | -7.8554E-01 | 2.3341E-02 |
| $\beta_3$ (H-H Combo) | -3.8316E-01 | -8.0779E-01 | 4.1743E-02 |
| $\beta_3$ (W-B/H Combo) | -4.4363E-01 | -8.1959E-01 | -6.6467E-02 |
| $\beta_3$ (W-W Combo) | -5.0121E-01 | -8.6441E-01 | -1.3658E-01 |
| $\beta_4$ (Casual Type) | 5.7433E-01 | 4.1160E-01 | 7.3812E-01 |
| $\beta_5$ (Duration x Casual Type) | 1.2236E-03 | 5.3148E-04 | 1.9334E-03 |
| $\beta_6$ (Combined Age) | -4.7206E-02 | -6.4325E-02 | -3.0012E-02 |
| $\beta_7$ (Combined Age$^2$) | 2.1339E-04 | 9.9824E-05 | 3.2592E-04 |
| $\beta_8$ (HIV+ Concordant) | -1.6064E+00 | -2.1535E+00 | -1.1241E+00 |
| $\beta_9$ (PrEP Use) | -5.2641E-01 | -6.8131E-01 | -3.7338E-01 |
| $\beta_{10}$ (San Francisco residence) | 5.4727E-02 | -2.9634E-01 | 3.9615E-01 |

Abbreviations: CI, confidence interval; B-H/W, black ego with either a Hispanic or white alter; H-B/W, Hispanic ego with either a black or white alter; H-H, Hispanic ego with a Hispanic alter; W-B/H, white ego with either a black or Hispanic alter; W-W, white ego with a white alter; PrEP, preexposure prophylaxis.

For the logistic regression model of one-time partnerships, we used a similar logistic regression approach as for persistent partnerships but dropped the partnership duration and partnership type (since there was only one type for this model) predictor variables. The corresponding linear model formula for persistent partnerships was as follows:

$$Y_i \sim \beta_0 + \beta_1 X_1 + \beta_2 X_2 + \beta_3 X_2^2 + \beta_4 X_3 + \beta_5 X_4 + \beta_6 X_5$$

where:

$Y_i$ = Log odds of the probability of condom use per one-time contact.

$X_1$ = Racial/ethnic combination of the ego (respondent) and alter (partner), coded in 6 categories to capture within and across group mixing: black-black, black-Hispanic/white, Hispanic-black/white, Hispanic-Hispanic, white-black/Hispanic, white-white.

$X_2$ = The combined age of ego and alter in years.

$X_3$ = The concordant diagnosed HIV-positive status of both ego and alter (1 = concordant positive; 0 = all other combinations of dyadic HIV status).

$X_4$ = Current use of pre-exposure prophylaxis (PrEP) by the ego (respondent) (1 = yes; 0 = no)

$X_5$ = Residence (1 = San Francisco; 0 = all other areas).

The coefficients for the model, and their lower and upper 95% confidence intervals, are presented in **Supplemental Table 7** below. Taking the inverse logit of the linear combination of coefficients will yield to the strata-specific predicted probabilities of condom use within the partnership.

**Supplemental Table 7.** Statistical Model of Per-Act Condom Use Probability for One-Time Sexual Contacts

| Model Parameter | Estimate | Lower 95% CI | Upper 95% CI |
|---|---|---|---|
| $\beta_0$ (Intercept) | 2.4591E+00 | 1.6900E+00 | 3.2311E+00 |
| $\beta_1$ (B-H/W Combo) | 1.5275E-01 | -3.7051E-01 | 6.7660E-01 |
| $\beta_1$ (H-B/W Combo) | -1.1786E-01 | -5.4023E-01 | 3.0441E-01 |
| $\beta_1$ (H-H Combo) | -1.5368E-01 | -6.0784E-01 | 3.0033E-01 |
| $\beta_1$ (W-B/H Combo) | -1.4286E-01 | -5.4014E-01 | 2.5453E-01 |
| $\beta_1$ (W-W Combo) | -2.6419E-01 | -6.5093E-01 | 1.2260E-01 |
| $\beta_2$ (Combined Age) | -5.4173E-02 | -7.3295E-02 | -3.5041E-02 |
| $\beta_2$ (Combined Age$^2$) | 2.6366E-04 | 1.3481E-04 | 3.9197E-04 |
| $\beta_3$ (HIV+ Concordant) | -1.8194E+00 | -2.6366E+00 | -1.1443E+00 |
| $\beta_4$ (PrEP Use) | -6.9183E-01 | -8.5210E-01 | -5.3351E-01 |
| $\beta_5$ (San Francisco residence) | -4.3426E-01 | -8.3380E-01 | -5.2430E-02 |

Abbreviations: CI, confidence interval; B-H/W, black ego with either a Hispanic or white alter; H-B/W, Hispanic ego with either a black or white alter; H-H, Hispanic ego with a Hispanic alter; W-B/H, white ego with either a black or Hispanic alter; W-W, white ego with a white alter; PrEP, preexposure prophylaxis.

##### *4.2.3 Predicted Probabilities in Epidemic Model*

Predicted probabilities of condom use conditional on an anal intercourse act were calculated based on the linear combination of partnership and individual attributes obtained dynamically by predicting from the statistical model with inputs based on the current simulated population. This set of predictors was input into a predict function in R to obtain the expected mean probabilities.

In Supplemental Figure 2 below, we display some example probabilities based on a subset of model inputs. This figure shows that condom use is lower in partnerships of a longer duration, higher in casual compared to main partnerships, higher when both partners are younger, and lower in partnerships in which the ego (respondent) reported currently using PrEP. Other predicted probabilities may be obtained from Supplemental Table 6 by taking the inverse logit of the linear combination of coefficients of interest.

Supplemental Figure 3 shows the predicted probabilities for the second logistic model, for condom use within one-time anal intercourse contacts. Here we display variation in condom use by combined age of the partners, current PrEP use, and racial combination of the partners. As the figure shows, condom use is higher within partners of a lower combined age, higher in partnerships involving black MSM (race.combo = 1 or 2), and lower among current PrEP users.

**Supplemental Figure 2.** Predicted Probabilities of Condom Use Per anal intercourse Act in Persistent Partnerships from the Logistic Regression Model, by Partnership Duration, Partnership Type (ptype: 1 = Main; 2 = Casual), Combined Partner Age (comb.age: 40 or 80 years), and PrEP Use.

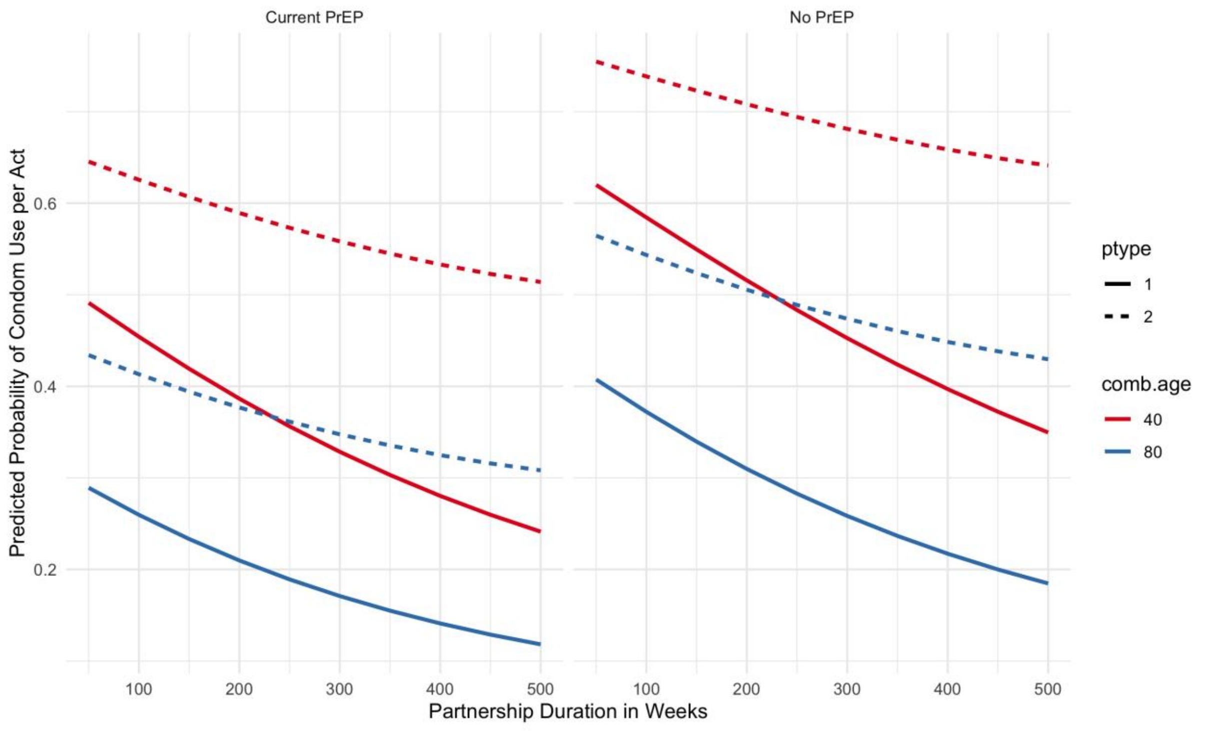

Based on these model predictions, which represent expected probabilities for each linear combination, we then drew individual probabilities of condom use per act in *EpiModel* using the rbinom function to draw randomly from the Bernoulli distribution with a vector of parameters, one value for each act. This generated a set of 0's and 1's for whether condom use occurred within the act as a function of the predictors in the statistical model.

**Supplemental Figure 3.** Predicted Probabilities of Condom Use in One-Time anal intercourse Contacts from the Logistic Regression Model, by Combined Partner Age, Current PrEP Use, and Racial Combination of Partners (race.combo: 1 = black ego-black alter; 2 = black ego-Hispanic or white alter; 3 = Hispanic ego-black or white alter; 4 = Hispanic ego-Hispanic alter; 5 = white ego-black or Hispanic alter; 6 = white ego-white alter.

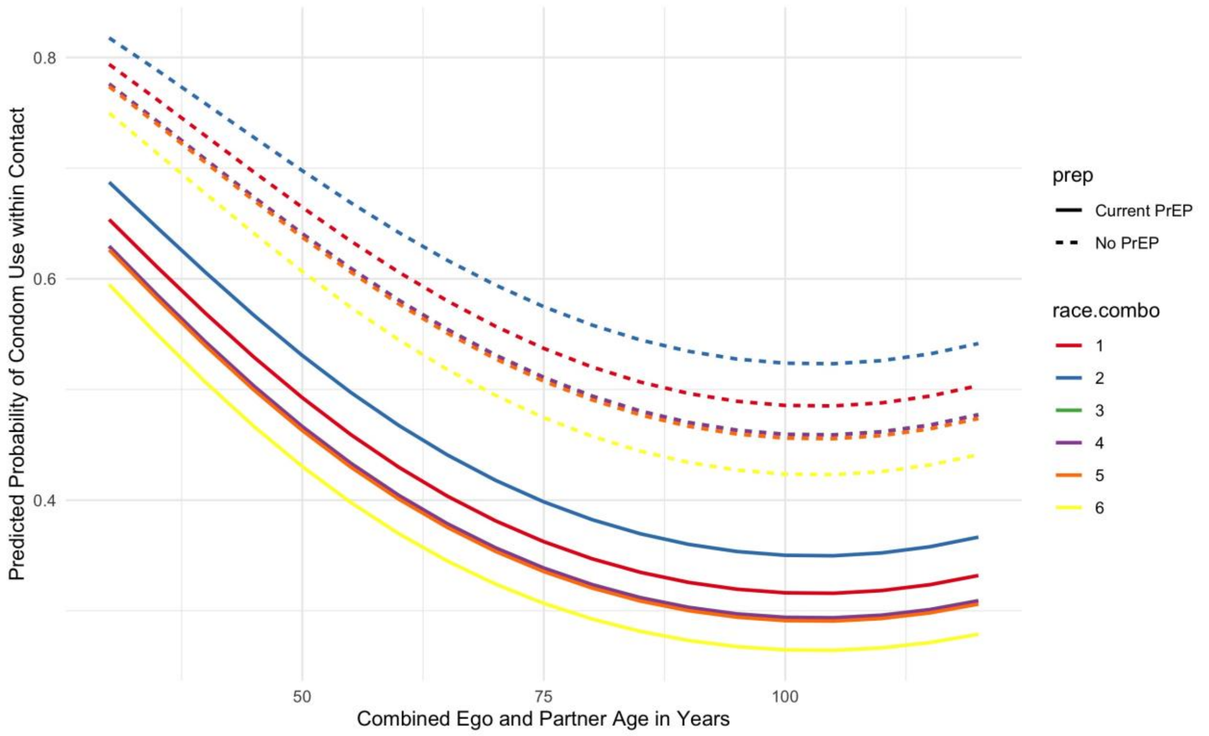

#### 4.4 Sexual Role

Men were assigned an individual sexual role preference (exclusively insertive, exclusively receptive, or versatile) as described in Section 3.1.9. Relationships between two exclusively insertive or two exclusively receptive men are prohibited via the TERGM models. Versatile men were further assigned a preference for being an the insertive partner drawn from a uniform distribution between 0 and 1 upon entry into the population; we refer to this proportion as the "insertivity quotient". When two versatile men are simulated to have an anal intercourse act, their sexual positions during that act must be determined (all other allowed combinations have only one direction). One option is for men to engage in intra-event versatility (IEV; i.e. both men engage in insertive and receptive anal intercourse during the act). The probability of this is was derived from the partner-specific role data described in Section 3.1.9. If IEV does not occur, then each man's probability of being the insertive partner equals his insertivity quotient divided by the sum of the two men's insertivity quotients.

### 5 DEMOGRAPHY AND INITIAL CONDITIONS

In this model, there are three demographic processes: entries, exits, and aging. Entries and exits are conceptualized as flows into and out of the sexually active population of interest: MSM aged 15 to 65 years old. Entry into this population represents the time at which persons become at risk of infection via male-to-male sexual intercourse, and we model these flows as starting at an age associated with sexual debut and ending at an age potentially before death (age 65). This age range also mapped directly on to the eligibility criteria of the ARTnet study.[8]

##### 5.1 Arrivals at Sexual Onset

All persons enter the network at age 15, which was the lower age boundary of ARTnet. The number of new entries at each time step was based on a fixed rate (0.045 per 100 person-weeks) that kept the overall network size in a relatively stable state. The model parameter governing this rate was tuned iteratively in order to generate simulations with a population size at equilibrium, given the inherent variability in population flows related to background mortality, sexual cessation (i.e., reaching the upper age limit of 65), and disease-induced mortality. At each time step, the exact number of men entering the population was simulated by drawing from a Poisson distribution with the rate parameter.

##### 5.2 Initialization of Attributes

Persons entering the population were assigned attributes in different categories. Some attributes remained fixed by definition (e.g., race/ethnicity), others were fixed by assumption (e.g., insertive versus receptive sexual role), and others were allowed to vary over time (e.g., age and disease status). Here we describe attributes initialized at the outset in the model and for arrivals into the population at each time step:

- **Race/ethnicity.** This model was based on a race/ethnic population composition categorized into three mutually exclusive groups: black, Hispanic, and white. At the outset of the model simulations, individuals were randomly assigned into one of these three groups with a probability equal to the proportions each represented in the Atlanta metropolitan target population based on 2018 Census data estimates for men aged 15 to 65. Those probabilities were: 3.8% black, 15.3% Hispanic, and 80.9% white. Incoming nodes during the dynamic simulation were also randomly assigned a race/ethnicity in these proportions.
- **Age.** In the dynamic simulation, as noted above, all incoming nodes were assigned an age of 15, which incrementally grew in weekly time steps. At the outset of the model simulations, we assigned nodes an age based on a uniform distribution, with ages from 15 to 65. This population-level age distribution was expected to converge to a more realistic distribution during model burn-in and calibration (explained in Section 9.2).
- **HIV Status.** In the dynamic simulation, all incoming nodes were assigned an HIV status of uninfected upon arrival into the population. This reflects the assumption that arrival corresponded with sexual debut, before which exposure to HIV would be very rare. At the outset of the model simulations, we randomly seeded the nodes with HIV infection by fitting and predicting from a logistic regression of diagnosed HIV status from the ARTnet data. This model incorporated city (residence in San Francisco), age, and race/ethnicity as the primary predictors based on the self-reported diagnosed HIV status reported by ARTnet respondents. These initial infections were all

assumed to be diagnosed based on this outcome. We did not expect that this initial condition of diagnosed HIV prevalence at the outset of the burn-in model to match the calibrated disease prevalence prior to experimental intervention models; instead this statistical modeling approach allowed for a data-driven seeding of HIV infection in the population that was distributed according to known demographic and geographic heterogeneity. Further description of the transition from initial HIV conditions to calibrated levels are provided in Section 8.2.

- **Circumcision Status.** Circumcision status was randomly assigned to incoming nodes at arrival and for all nodes as initial conditions in the simulations. Based on empirical data from San Francisco MSM,[18] 62% of black men were circumcised before sexual onset, 42% of Hispanic men, and 55% of white men. As described in Section 8, circumcision was associated with a 60% reduction in the per-act probability of infection for HIV-negative males for insertive anal intercourse only (i.e., circumcision did not lower the *transmission* probability if the HIV-positive partner was insertive).[2,19]

##### 5.3 Departures from the Network

All persons exited the network by age 65, either from mortality or by reaching the upper age bound of the MSM target population of interest. This upper limit of 65 was modeled deterministically (probability = 1), but other exits due to mortality were modeled stochastically. Departures included both natural (non-HIV) and disease-induced mortality causes before age 65. Background mortality rates were based on US all-cause mortality rates specific to age and race/ethnicity from the National Vital Statistics life tables.[20] Note that these rates include deaths due to HIV/AIDS; however, the relative fraction of those deaths to total deaths is small enough not to impact this background mortality process. **Supplemental Table 8** shows the probability of mortality per year by age and race/ethnicity.

**Supplemental Table 8.** Age- and Race/Ethnicity-Specific Probabilities of Mortality among Men in the United States

| Age | Black | Hispanic | White |
|---|---|---|---|
| 15–19 | 0.00124 | 0.00062 | 0.00064 |
| 20–24 | 0.00213 | 0.00114 | 0.00128 |
| 25–29 | 0.00252 | 0.00127 | 0.00166 |
| 30–34 | 0.00286 | 0.00132 | 0.00199 |
| 35–39 | 0.00349 | 0.00154 | 0.00226 |
| 40–44 | 0.00422 | 0.00186 | 0.00272 |
| 45–49 | 0.00578 | 0.00271 | 0.00382 |
| 50–54 | 0.00870 | 0.00440 | 0.00591 |
| 55–59 | 0.01366 | 0.00643 | 0.00889 |
| 60–64 | 0.02052 | 0.00980 | 0.01266 |

These yearly probabilities were transformed into weekly risks. Natural mortality was then applied to persons within the population at each time step stochastically by drawing from a Bernoulli distribution for each eligible person with a probability parameter corresponding to their age- and race-specific risk of death. Disease-related mortality, in contrast, was modeled based on clinical disease progression, as described in Section 6.

##### 5.4 Aging

The aging process in the population was linear by time step for all persons. The unit of time step in these simulations was one week, and therefore, persons were aged in weekly steps between the minimum and maximum ages allowed (15 and 65 years old). Evolving age impacted background mortality, age-based mixing in forming new partnerships, and other features of the epidemic model described below. Persons who exited the network were no longer active and their attributes such as age were no longer updated.

#### 6 INTRAHOST EPIDEMIOLOGY

Intrahost epidemiology includes features related to the natural disease progression within HIV-positive persons in the absence of clinical intervention. The main component of progression that was explicitly modeled for this study was HIV viral load. In contrast to other modeling studies that model both CD4 and viral load, our study used viral load progression to control both interhost epidemiology (HIV transmission rates) and disease progression eventually leading to mortality.

Following prior approaches,[1,2,4,6,21] we modeled changes in HIV viral load to account for the heightened viremia during acute-stage infection, viral set point during the long chronic stage of infection, and subsequent rise of viral load at clinical AIDS towards disease-related mortality. The HIV viral load has a direct impact on the rates of HIV transmission within serodiscordant pairs in the model, and this interaction is detailed in Section 8. A starting viral load of 0 is assigned to all persons upon infection. From there, the natural viral load curve is fit with the parameters described in **Supplemental Table 9**.

| **Supplemental Table 9.** HIV Natural History Parameters | | |
|---|---|---|
| **Parameter** | **Value** | **Reference** |
| Time to peak viremia in acute stage | 45 days | Little[22] |
| Level of peak viremia | 6.886 $log_{10}$ | Little[22] |
| Time from peak viremia to viral set point | 45 days | Little,[22] Leynaert[23] |
| Level of viral set point | 4.5 $log_{10}$ | Little[22] |
| Duration of chronic stage infection (no ART) | 3550 days | Buchbinder,[24] Katz[25] |
| Duration of AIDS stage | 728 days | Buchbinder[24] |
| Peak viral load during AIDS | 7 $log_{10}$ | Estimated from average duration of AIDS |

After infection, it takes 45 days to reach peak viremia, at a level of 6.886 $log_{10}$. From peak viremia, it takes another 45 days to reach viral set point, which is set at a level of 4.5 $log_{10}$. Changes occur linearly on the log scale. The total time of acute stage infection is therefore 3 months. The duration of chronic stage infection in the absence of clinical intervention is 3550 days, or 9.7 years. The total duration of pre-AIDS disease from infection is therefore approximately 10 years. At onset of AIDS, HIV viral load rises linearly on the log scale from 4.5 $log_{10}$ to 7 $log_{10}$. The time spent in the AIDS stage is 728 days, or 2 years. This viral load trajectory is for ART-naïve persons only, and the influence of ART on disease progression is detailed in Section 7. These transitions are deterministic for all ART-naïve persons. In the AIDS stage, disease-related mortality is imposed stochastically with a homogenous risk of 1/104, corresponding to average duration of the AIDS stage in weeks. This is accomplished by drawing from a binomial (Bernoulli) distribution for all eligible individuals in the AIDS stage.

#### 7 CLINICAL EPIDEMIOLOGY

Clinical epidemiological processes in the model refer to all steps along the HIV care continuum after initial HIV infection: diagnosis, linkage to ART care, adherence to ART, and HIV viral load suppression. In this model, these clinical features have interactions with the behavioral features detailed above, as well as impacts on the rates of HIV transmission, detailed in the next section. The features of our model's clinical processes generally follow the steps of the HIV care continuum, in which persons transition across states from infection to diagnosis to ART initiation to HIV viral suppression.[26]

##### 7.1 HIV Diagnostic Screening

Both HIV-uninfected and HIV-infected persons in our model were exposed to regular interval-based HIV screening that served as a common entry point for HIV prevention and HIV treatment services, respectively. Individuals screened at routine intervals first based on whether they were currently using PrEP or not.

Our approach to parameterization for HIV screening among PrEP non-users was first to start with priors based on ARTnet data for time since last HIV test for HIV-uninfected, and then use model calibration (the technical details of which are explained in Section 9) to fit these parameters to reproduce the level of the first step of the HIV care continuum (the fraction of HIV-infected persons who were diagnosed). For this we have used general population values as the ones specific to MSM and stratified by race were not available in San Francisco's report[27]. We used that approach because self-reported HIV screening data alone may be biased, and this calibration approach allows for triangulation of diagnostic history based on more objective data.

The three model parameters for the weekly screening rates were calibrated to meet the target statistics, which were the fraction of HIV-infected MSM who were diagnosed. After the calibration, the model was simulated 20,000 times and the numerical results from this parameterization are shown in Supplemental Table 10.

**Supplemental Table 10.** Model Parameterization for HIV Screening

| | **Black MSM** | **Hispanic MSM** | **White MSM** |
|---|---|---|---|
| Target Statistic: Diagnosed Fraction[28] | 93% | 93% | 93% |
| Simulations: Diagnosed Fractions | 92.8% [91.1%, 94.4%] | 92.8% [91.8%, 93.7%] | 92.7% [92.2%, 93.2%] |
| Calibrated Rates (per Week) | 0.007 | 0.007 | 0.007 |

Abbreviation: MSM, men who have sex with men.
Results as: Median [1st quartile, 3rd quartile]

The target statistics for the diagnosed fraction were drawn from the 2014 San Francisco Department of Public Health surveillance report based data for MSM in 2013[27] (the technical details of which are explained in Section 9). After calibration, the simulated diagnosed fractions were nearly identical to those targets.

Supplemental Figure 4 shows the general results of this calibration. The model starts with all persons with HIV infection as undiagnosed, then the model is simulated for 60 years (x-axis time scale is in weeks) to establish stable equilibrium conditions for this and the other calibrated parameters. The target statistics are shown with dashed horizontal lines and the simulated statistics are shown with solid lines.

For PrEP users, we modeled HIV screening practice based on CDC clinical practice guidelines.[29] The guidelines recommend ongoing screening at 3-month intervals for MSM actively using PrEP. This schedule was imposed for all PrEP users active in their PrEP use, regardless of PrEP adherence categories. We also assumed no racial/ethnic variation in HIV screening rates for PrEP users.

Finally, we also modeled a 21-day window period after infection during which the tests of the truly HIV+ persons would show as negative to account for the lack of antibody response immediately after infection.[30] HIV+ persons who tested after this window period would be correctly diagnosed with 100% test sensitivity. MSM with recent but undetected infection were still eligible for PrEP initiation since PrEP eligibility was based diagnosed HIV status. This would have resulted in a period in which HIV-infected but undiagnosed persons were classified as on PrEP. This did not impact their HIV transmission potential (and could not impact their acquisition potential). This undetected infection would then be identified at the next quarterly PrEP clinical visit, at which point they would be transitioned off PrEP.

##### 7.2 Antiretroviral Therapy (ART) Initiation

Following HIV diagnosis, individuals were linked to HIV care that provided ART. In the absence of quantitative data and based on current clinical practice guidelines for MSM in the U.S., we assumed no gap between treatment entry and ART initiation. Although the intermediate steps of the HIV care continuum are often characterized by any linkage to HIV care and/or ART, we selected a second HIV care continuum target of linkage to HIV care specifically within three months of diagnosis for two reasons. First, in the dynamic modeling context, the temporally defined threshold easily mapped on to the tracking implemented for simulated individuals in the model. Second, there were readily available surveillance estimates for this outcome. With respect to the latter, we used data from the San Francisco Department of Public Health care continuum estimates for 2013, stratified by transmission risk level and race/ethnicity. We assume therefore that there is an exponential statistical relationship between the proportion linked to care within three months and the average time to care entry following diagnosis. This time-to-event estimate below is generally consistent with recent cohort data that suggest relatively rapid ART initiation following diagnosis.[31]

The approach was similar to calibration for HIV screening rates. Over the 65-year burn-in simulation period, persons were linked to HIV care with ART with initiation rates that were specific to race/ethnicity. The specific metric used within the simulations to compare against the target statistics was the time period between diagnosis and first ART use, which were uniquely tracked for all individuals with HIV infection in the model. A group-specific proportion of persons whose difference between diagnosis and ART initiation was less than or equal to four weeks was calculated in the model. The target statistics are shown with dashed horizontal lines and the simulated statistics are shown with solid lines.

After the calibration, the model was simulated 20,000 times and the numerical results from this parameterization are shown in Supplemental Table 11. The rate of care establishment was highest for white MSM, and lower for Hispanic and mostly for black MSM. With the calibrated rates, the model simulations matched these target statistics. The inverse of these rates implied that the average time to ART initiation after HIV diagnosis was between 6 to 8 weeks on average.

| **Supplemental Table 11.** Model Parameterization for ART Linkage After Diagnosis | | | |
|---|---|---|---|
| | **Black MSM** | **Hispanic MSM** | **White MSM** |
| Target Statistic:<br>Fraction Linked within 3m[27] | 82% | 89% | 90% |
| Simulations: Fraction Linked | 84.4%<br>[82.0%, 86.6%] | 90.7%<br>[89.6%, 91.6%] | 91.5%<br>[91.1%, 91.9%] |
| Calibrated Rates (per Week) | 0.125 | 0.158 | 0.164 |

Abbreviations: ART, antiretroviral therapy; m, month; MSM, men who have sex with men.
Results as: Median [1st quartile, 3rd quartile]

Supplemental Figure 4 shows the general results of this calibration. The model starts with all persons with HIV infection as undiagnosed, then the model is simulated for 65 years to establish stable equilibrium conditions for this and the other calibrated parameters. The target statistics are shown with dashed horizontal lines. The medians of simulated statistics are shown with solid lines surrounded by a ribbon representing the Inter Quartile Range (IQR)

**Supplemental Figure 4.** Fraction of MSM with HIV Who Are Linked to Care Within 3 Months, Simulations versus Target Statistics, Stratified by Race/Ethnicity (red = black MSM, green = Hispanic MSM, Blue = white MSM)

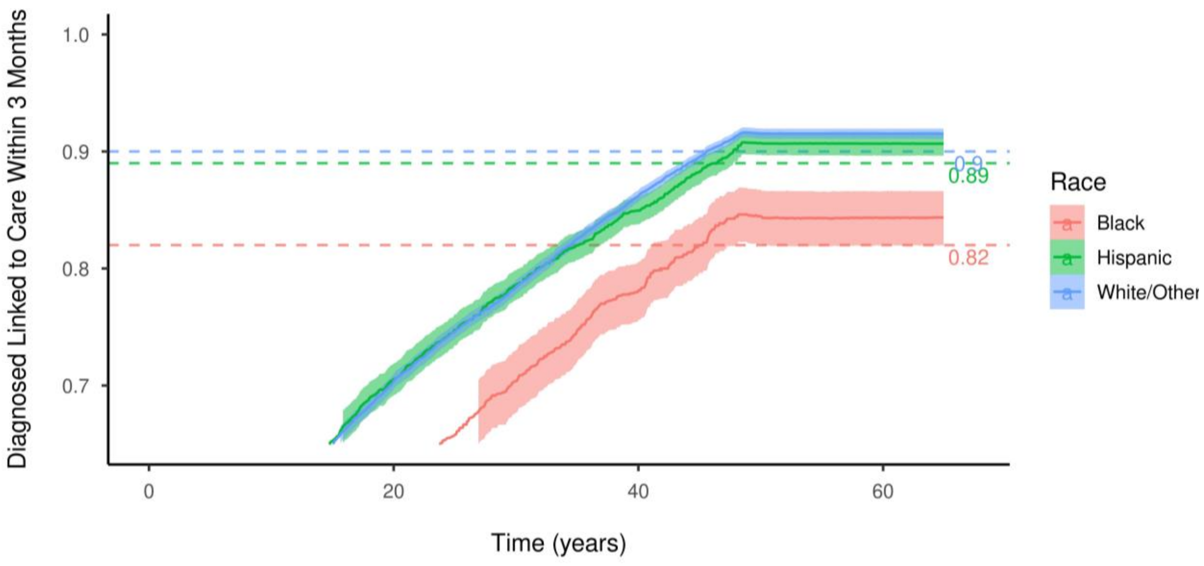

#### 7.3 ART Adherence and HIV Viral Load Suppression

MSM who initiated ART could cycle on and off treatment, where cycling off treatment resulted in an increase in the viral load back up to the assumed set point of 4.5 $\log_{10}$. The slope of changes to viral load were calculated such that it took a total of 3 months to transition between the set point and the on-treatment viral loads.[32] Individuals on ART could reach full suppression with sustained ART use. The nadir HIV viral load level was assumed to be 1.5 $\log_{10}$ among those at full suppression levels.[32] The latter corresponds to an absolute viral load below the standard levels of detection (viral load = 50).[33] Viral load was tracked and updated continuously over time based on the natural history of HIV disease by stage, and current use of ART.

The patterns of ART adherence (cycling on and off ART) leading to full HIV viral suppression were estimated based on an analysis of HIV care patterns among MSM in San Francisco[27] and model calibration similar to the first two HIV care continuum steps. The rates of cycling off ART after initially starting (the “halting rate”), the rates of cycling back on after a period of stopping (the “reinitiation rate”) and the fraction of the population able to reach viral suppression (“Partial Suppression Fraction”) controlled overall levels of HIV viral suppression (viral load < 50 among the treated) as well as the levels of durable HIV viral suppression (at least one year of viral suppression among the virally suppressed). The “Partial Suppression Fraction” correspond to the individuals treated by ART but unable to reach viral load < 50. This could depict situation where individuals are currently on treatment but do not take it correctly for example.

After the calibration, the model was simulated 20,000 times and the numerical results from this parameterization are shown in Supplemental Table 12.

**Supplemental Table 12.** Model Parameterization for ART Retention Rates After Linkage

| | **Black MSM** | **Hispanic MSM** | **White MSM** |
|---|---|---|---|
| Target Statistic: Treated Fraction [27] | 82% | 86% | 88% |
| Simulation: Treated Fraction [27] | 84.3%<br>[81.9%, 86.6%] | 86.1%<br>[84.9%, 87.3%] | 88.0%<br>[87.5%, 88.5%] |
| Target Statistic: Suppressed Fraction[27] | 71% | 71% | 71% |
| Simulation: Suppressed Fraction[27] | 70.0%<br>[66.7%, 73.3%] | 70.2%<br>[68.5%, 72.0%] | 73.2%<br>[72.3%, 74.0%] |
| Target Statistic:<br>Durably Suppressed Fraction [27] | 92% | 92% | 92% |
| Simulation: Durably Suppressed Fraction [27] | 92.1%<br>[89.6%, 94.2%] | 91.8%<br>[90.5%, 93.0%] | 91.7%<br>[91.1%, 92.2%] |
| Calibrated: Partial Suppression Fraction | 0.4730 | 0.4730 | 0.4325 |
| Calibrated: Stopping Rates (per week) | 0.0009 | 0.0009 | 0.00089 |
| Calibrated: Cycling Back Rates (per week) | 0.00377 | 0.0046 | 0.0057 |

Abbreviations: ART, antiretroviral therapy; MSM, men who have sex with men; viral load, viral load.
Results as: Median [1st quartile, 3rd quartile]

Supplemental Figure 5 and 6 shows the general results of this calibration. The model starts with all persons with HIV infection as undiagnosed, then the model is simulated for 65 years to establish stable equilibrium conditions for this and the other calibrated parameters. The target statistics are shown with dashed horizontal lines. The medians of simulated statistics are shown with solid lines surrounded by a ribbon representing the Inter Quartile Range (IQR)

**Supplemental Figure 5.** Fraction of MSM Diagnosed With HIV Who Are Treated With ART, Simulations Versus Target Statistics, Stratified By Race/Ethnicity (Red = Black MSM, Green = Hispanic MSM, Blue = White MSM)

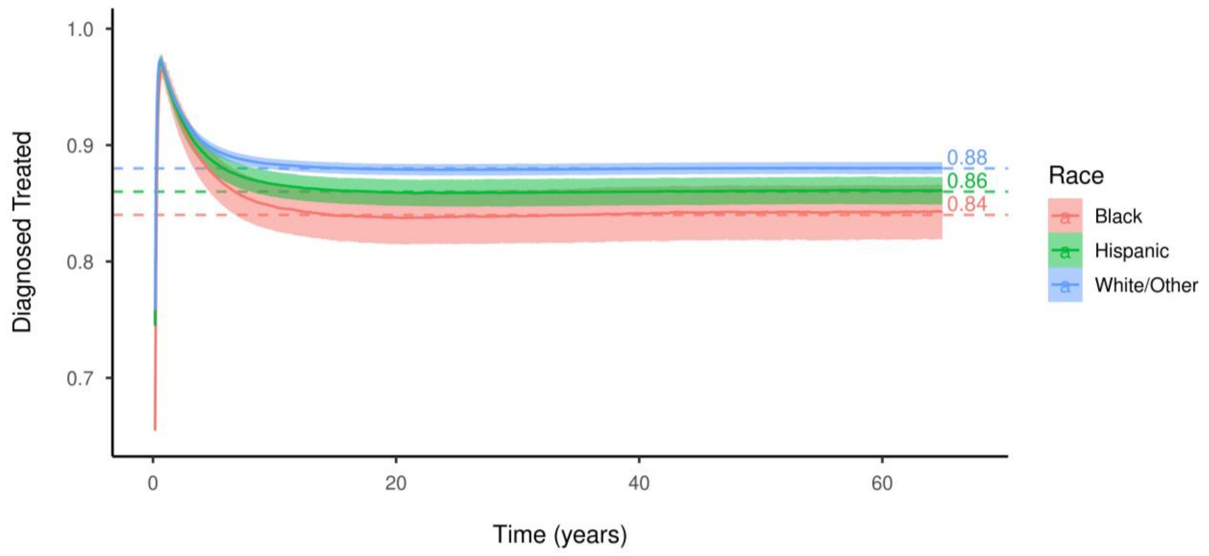

**Supplemental Figure 6.** Fraction of MSM With HIV And Treated by ART Who Are Virally Suppressed, Simulations Versus Target Statistics, Stratified By Race/Ethnicity (Red = Black MSM, Green = Hispanic MSM, Blue = White MSM)

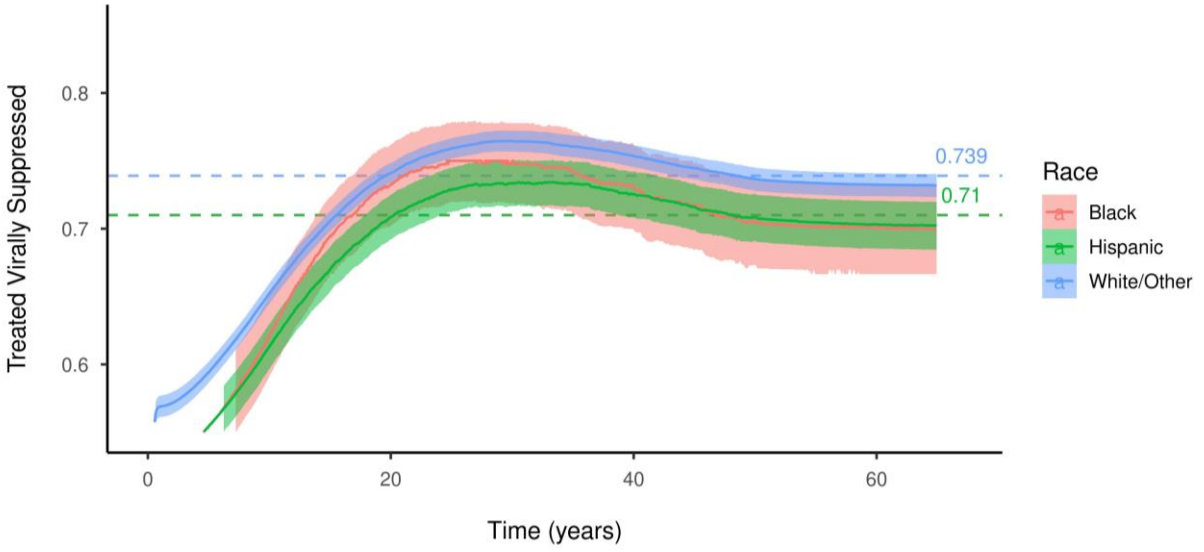

The corresponding halting rates were therefore slightly lower in white/other MSM compared to the other two groups. The inverse of these rates implied a time to first stopping ART after initiation of 21 years, consistent with the high prevalence but relatively low incidence of HIV in San Francisco.

#### 7.4 AIDS Disease Progression and AIDS-Related Mortality

Progression to AIDS after ART initiation was modeled based on the cumulative time on and off ART for individuals who had been linked to treatment (persons never linked to ART progressed according to the rates in Section 6). The maximum untreated time between infection and the start of AIDS was 9.7 years.[24] We assumed that an individual who had once initiated ART could spend a maximum of 15 years off of ART over the life course before progression to AIDS, similar to previous models.[1] Persons who had ever initiated ART progressed through AIDS at a similar rate as those who were ART-naïve, but ART use during the AIDS stage was associated with the same declines in HIV viral load as in pre-AIDS stages. However, to account for treatment failure during the AIDS stage, the same mortality rate was applied to persons on active ART and those not on active ART within the AIDS stage. Therefore, we assumed that the probability of disease-induced mortality given AIDS was 1/104 weeks, consistent with approximately 2 years on average spent in the AIDS stage during untreated infection.

#### 7.5 PrEP Initiation and Adherence

PrEP initiation and retention were modeled based on the proportion of MSM who tried PrEP in the past 12 months and the median time people stay on PrEP. In our models, we consider that PrEP initiation can only occur after a negative HIV test. This makes the PrEP initiation rate linked to the test rate. PrEP start and stop rate are thus calibrated after the other parameters (the technical details of which are explained in Section 9.2).

After the calibration, the model was simulated 560 times and the numerical results from this parameterization are shown in Supplemental Table 13.

**Supplemental Table 13.** Model Parameterization for PrEP starting and stopping rate

| | **All MSM** |
|---|---|
| Target Statistic: Any PrEP 12m[34] | 45% |
| Simulation: Any PrEP 12m | 42.6% [42.9%, 43.3%] |
| Target Statistic: Median Time on PrEP (weeks)[34] | 25 |
| Simulation: Median Time on PrEP (weeks) | 24 [24, 25] |
| Calibrated: PrEP Starting Rate (on negative screening) | 68.5% |
| Calibrated: PrEP Stopping Rate (per week) | 0.0259 |

Using these parameters yielded a PrEP coverage (proportion of users among the eligible) of 25%.

### 8 INTERHOST EPIDEMIOLOGY

Interhost epidemiological processes represent the HIV-1 disease transmission within the model. Disease transmission occurs between sexual partners who are active on a given time step. This section will describe how the overall rate is calculated as a function of the intrahost epidemiological profile of each member of a partnership, and behavioral features within the dyad.

#### 8.1 HIV-Discordant Dyads

At each time step in the simulation, a list of active dyads was selected based on the current composition of the network. This was called an "edgelist." Given the three types of partnerships detailed above, the full edgelist was a concatenation of the type-specific sublists. The complete edgelist reflects the work of the STERGM- and ERGM-based network simulations, wherein partnerships formed on the basis of nodal attributes and degree distributions (see Section 3). From the full edgelist, a disease-discordant subset was created by removing those dyads in which both members were HIV-negative or both were HIV-positive. This left dyads that were discordant with respect to HIV status, which was the set of potential partnerships over which infection may be transmitted at that time step.

#### 8.2 HIV Transmission Rates

Within HIV-discordant dyads, transmission was simulated stochastically across separate sexual acts at each timestep. The per-act probabilities were a combined function of attributes of the HIV-negative and HIV-positive partner. These probabilities were calibrated to reach the empirical diagnosed HIV prevalence. The final per-partnership transmission rates per time step were then a function of these one minus per-act transmission probabilities raised to the number of acts within the partnership during that time step.

##### *8.2.1 Per-Act Transmission Probabilities*

Within disease-discordant dyads, HIV transmission was modeled based on a sexual act-by-act basis, in which multiple acts of varying infectiousness could occur within one partnership within a weekly time step. Determination of the number of acts within each discordant dyad for the time step, as well as condom use and role for each of those acts, was described in Section 4. Transmission by act was then modeled as a stochastic process for each discordant sex act following a Bernoulli distribution with a probability parameter that is a multiplicative function of the following predictors of the HIV-negative and HIV-positive partners within the dyad, as shown in Supplemental Table 14 below.

For each act, the overall transmission probability was determined first based on sexual position and HIV viral suppression status of the infected partner. If the infected partner was virally suppressed and on ART, then the base probability was 2.2/100,000, which was derived from a model-based estimate of Supervie.[35] This study estimated the upper bound of the transmission probability of 4.4/100,000 for MSM; we used the mean between the observed number (zero) and this upper bound as our base per-act transmission probability (so 2.2 transmissions per 100,000 exposures) in our model.

If the infected partner was not virally suppressed (at conditions of 200 copies/mL or higher) or not currently on ART, the base probability was a function of whether the HIV-negative partner was in the receptive or insertive role, with the former at a 2.6-fold infection risk compared to the latter. Then, following the parametric function of Wilson,[36] the HIV-positive partner's viral load modifies this base probability in a non-linear formulation, upwards if the viral load was above the viral load set point during chronic stage infection in the absence of ART, and downwards if it was below the set point.

Following others, we modeled an excess transmission risk in the acute stage of infection above that predicted by the heightened viral load during that period.[37] Three covariates could reduce the risk of infection: condom use within the act by either the HIV-negative or HIV-positive partner, circumcision status of the HIV- partner (only if the HIV- partner was insertive in that act), and PrEP use at the time of the act by the HIV- partner.

**Supplemental Table 14.** Per-Act Transmission Probabilities and Modifiers

| Predictor | Partner | Parameters | References |
|---|---|---|---|
| Sexual role (insertive or receptive) | HIV- | *Receptive:* 0.008938 base probability when HIV+ partner has 4.5 $\log_{10}$ viral load | Vittinghoff[38] |
| | | *Insertive:* 0.003379 base probability when HIV+ partner has 4.5 $\log_{10}$ viral load | Vittinghoff[38] |
| HIV viral load (viral load) | HIV+ (Not virally suppressed or not on ART) | Multiplier of $2.45^{(\text{viral load - 4.5})}$ on sexual-role specific base probabilities above | Wilson[36] |
| | HIV+ (Virally suppressed and on ART) | 2.2/100,000 base probability, regardless of sexual role | Supervie[35] |
| Acute stage | HIV+ | Multiplier of 6 | Leynaert,[23] Bellan[37] |
| Condom use | Both | Multiplier of 0.05 plus 0.25 | Varghese,[39] Weller,[40] Smith[41] |
| Circumcision status | HIV-, insertive | Multiplier of 0.40 | Gray[19] |
| Preexposure Prophylaxis (PrEP) | HIV- | High adherence: Multiplier of 0.01<br>Medium adherence: Multiplier of 0.19<br>Low adherence: Multiplier of 0.69 | Grant[42] |

For condom use, we updated our previous approach to explicitly represent condom failure that would result in a transmission event. Our previous models used estimates of HIV incidence comparing consistent condom users to occasional or non-condom users, resulting in a condom "efficacy" of 75–80%. However, this efficacy gap of 20–25% is the function of both the biological/physiological gaps in protection given perfect and consistent condom use during anal intercourse as well as the human error resulting in impact use. Such error could represent condom breakage, misapplication, incomplete use during sexual activity, and other related causes.[41] For this model, we assumed a 95% efficacy for the former, and a 25% absolute reduction in that efficacy as a function of condom failure to arrive at the previous range of 71% total effectiveness.

###### 8.2.2 Calibration of Transmission Probabilities

In addition to the calibration of the HIV care continuum parameters described in Section 7, we also calibrated the per-act transmission probabilities so that the diagnosed HIV prevalence was consistent with empirical data on HIV burden in this target population. Our target statistic for this calibration step was the estimated HIV prevalence by race/ethnicity. We used 25% as the overall HIV prevalence in the MSM population as the 2011 estimates for San Francisco were 22.7% for MSM and 46% for MSM who inject drugs[43]. We then used the proportion of each race/ethnicity from the San Francisco Department of public health in 2013 to calculate the prevalence. The target statistics of diagnosed HIV prevalence for MSM in San Francisco were 31.8% for black MSM, 27.3% for Hispanic MSM, and 24.0% for white MSM.

The per-act transmission probabilities defined above were then multiplied by a factor unique to each race/ethnic group. These calibration factors represent the additional sources of potential error in the transmission parameters that would generate the current HIV epidemic. These include co-factors not included in this model, such as untreated sexually transmitted infections.[44] The upweighting of the transmission probabilities for black MSM and down-weighting for white and Hispanic MSM is due to the long-standing finding that race-stratified behavioral and network data do not, by themselves, explain the excess burden of HIV among black MSM.[45,46]

After the calibration, the model was simulated 20,000 times and the numerical results from this parameterization are shown in Supplemental Table 15.

**Supplemental Table 15.** Model Parameterization for ART Retention Rates After Linkage

| | **Black MSM** | **Hispanic MSM** | **White MSM** |
|---|---|---|---|
| Target Statistic: HIV Prevalence[27,43] | 31.8% | 27.3% | 24.0% |
| Simulation: Treated Fraction [27] | 32.6% [30.7%, 34.4%] | 26.8% [25.7%, 27.8%] | 23.9% [23.2%, 24.6%] |
| Calibrated: Partial Suppression Fraction | 1.4 | 0.85 | 0.73 |

Abbreviations: ART, antiretroviral therapy; MSM, men who have sex with men; viral load, viral load.
Results as: Median [1st quartile, 3rd quartile]

Supplemental Figure 7 shows the general results of this calibration. The model starts with all persons with HIV infection as undiagnosed, then the model is simulated for 60 years (x-axis time scale is in weeks) to establish stable equilibrium conditions for this and the other calibrated parameters. The target statistics are shown with dashed horizontal lines and the simulated statistics are shown with solid lines.

**Supplemental Figure 7.** Prevalence of HIV in the Simulated population, Simulations versus Target Statistics, Stratified by Race/Ethnicity (Red = Black MSM, Green = Hispanic MSM, Blue = White MSM)

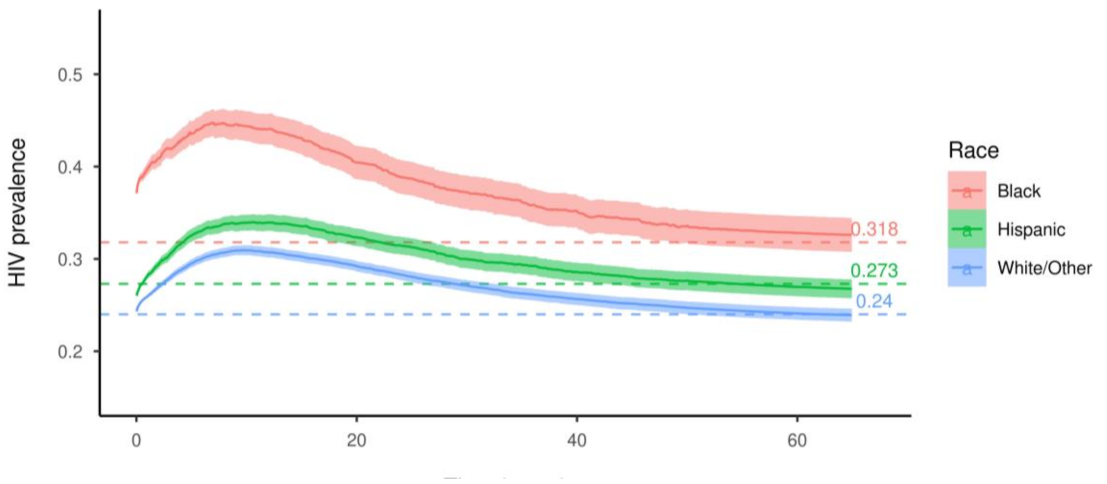

##### 8.2.3 *Final Per-Partnership-Week Transmission Rates*

The final transmission rate per partnership per weekly time step was a function of the per-act probability of transmission in each act and the number of acts per time step. The per-act transmission probability could be heterogeneous within a partnership due to various types of acts in each interval: for example, a HIV-negative man who is versatile in role may have both insertive and receptive intercourse within a single partnership; some acts within a partnership may be protected by condom use while others are condomless. Transmission was simulated for each act within each serodiscordant dyad, based on draws from a Bernouli distribution with the probability parameter equal to the per-act transmission probabilities detailed above.

### 9 MODEL CALIBRATION

This section describes the methods for executing the simulations and conducting the data analysis on the outcomes in further detail.

#### 9.1 Calibration Methods

We first used Bayesian approaches to define model parameters with uncertain values, construct prior distributions for those parameters, and fit the model to HIV/STI prevalence and incidence data to estimate the posterior distributions of those parameter values.

We used approximate Bayesian computation with sequential Monte Carlo sampling (ABC-SMC) methods[37,47] to calibrate behavioral parameters in which there was measurement uncertainty in order to match the simulated HIV prevalence at the end of the burn-in simulations to the targeted HIV prevalence.

The details of ABC depend on the specific algorithm used, but in this case, ABC-SMC proceeded as follows.

For each candidate parameter, $\theta$, to be estimated, we:

1. Sampled a candidate $\theta^i$ from a prior distribution $\pi(\theta)$
2. Simulated the epidemic model with candidate value, $\theta^i$.
3. Tested if a distance statistic, $d$ (e.g., the difference between observed HIV prevalence and model simulated prevalence) was greater than a tolerance threshold, $\epsilon$.
    a. If $d > \epsilon$ then discard
    b. If $d < \epsilon$ then add the candidate $\theta^i$ to the posterior distribution of $\theta$.
4. Sample the next sequential candidate, $\theta^{i+1}$, either independently from $\pi(\theta)$ (if 3a) or from $\theta^i$ plus a perturbation kernel with a weight based on the current posterior distribution (if 3b).

When sufficiently good posterior distribution for our parameters were obtained, we manually selected the values that yielded the outcomes closest to our targets.

#### 9.2 Calibration Steps

We took a two-step approach to implementing the model calibration. First, we calibrated the model to match the target statistics for the HIV care continuum (screening, linkage, and HIV viral load suppression) and diagnosed HIV prevalence in San Francisco in 2013, prior to the introduction of PrEP. This involved simulating the model at least 500 times for 65 years (the first burn-in period) and evaluating the distance between the selected target statistics and the simulations at the final year of the period. Once that calibration was complete, we simulated 20,000 replicates of the fitted model and selected the single simulation with the values of the target statistics closest to the targets (with total absolute deviance).

Second, we then simulated the model for an additional 4 years (representing the period between 2013 and 2017) in which PrEP was initially scaled up. The goal of this second burn-in period was to have PrEP coverage (the fraction of eligible MSM who currently use PrEP) calibrated to be approximately 25%. We accomplished this calibration by iteratively adjusting the model parameters for the probability of starting PrEP conditional on eligibility such that the final median PrEP coverage matched this target statistic. For this study, we had to perform this calibration step twice, once for the model scenarios in which it was assumed that PrEP initiation required an HIV-negative screening result, and another for the scenario that assumed that PrEP initiation was random (i.e., it did not require linkage to an HIV-negative screening event). The calibrated probability for the PrEP-linked scenario was 66.0% and the calibrated probability for the PrEP-unlinked scenario was 0.0411%. The probabilities are so different because the opportunities to start PrEP in the unlinked scenario were much higher than in the linked scenario, the latter of which require concurrent indications and an HIV-negative screening event.

#### 10 INTERVENTION SCENARIO PARAMETERS

Within the intervention component of the model, changes to ART coverage corresponded to changes in the halting rates by relative amounts compared to the base calibrates rates. Similarly, changes to PrEP coverage corresponded to changes in the halting rate by relative amounts compared to the base calibrates rate. We did not model a distinct clinical typology of ART users with a lower propensity for ART discontinuation, above and beyond the differences by race/ethnicity and not at all for PrEP, for two reasons. First, the empirical data to support a distinct typology at the population-level are insufficient. Second, the interventions currently in the scenarios are designed to shift the overall population averages rather than focus on a subgroup who would be at higher risk of ART or PrEP dropout.

##### 10.1 Primary Scenarios

As the calibrated PrEP coverage was 25% in San Francisco in 2017, we decided to use 12.5% increments, from 25% to 75% as counterfactual targets. We calibrated the corresponding stopping rates by adding four more years with the new parameter value to the second step of calibration (see Appendix 9.2). Supplemental Figure 8 below shows how the PrEP coverage changes over time in these scenarios.

**Supplemental Figure 8.** Evolution of the median PrEP coverage over the course of the Primary Scenarios. The “Initial PrEP ramp up” period correspond to the 2013-2017 period where PrEP was first introduced. The calibrated PrEP stopping rate is used there. The “Intervention period” correspond to the 10 years of intervention where the PrEP stopping rate is modified to get the new target coverage at year 8.

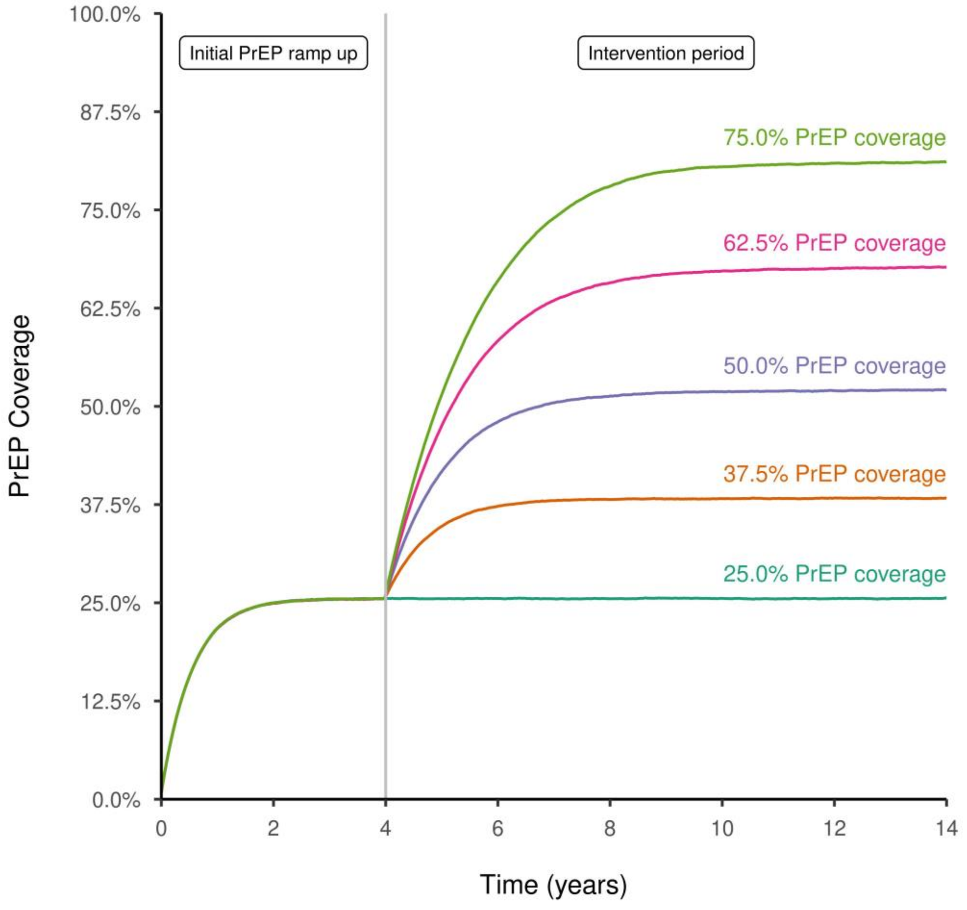

#### 10.2 Secondary Scenarios

In these scenarios, we use 3 different baseline ART coverage. We used the proportion of HIV virally suppressed MSM among all the infected as our calibration target. In the calibrated model, the value was 61%. We decided to calibrate the Low ART population to 50%, which means multiplying the ART stopping rates by 2.4. For the High ART population, we reduced the stopping rates to 0, meaning that no ART dropout was possible. It resulted in a 68% of HIV virally suppressed. The fact that it does not go higher is because we kept the fact that some MSM treated with ART would never reach HIV viral suppression. (See Appendix 7.3). For each one of the three ART population, we varied PrEP coverage. Again, we used 12.5% increments but from 0% to 75% as counterfactual targets. Supplemental Figure 9 below shows how the PrEP coverage changes over time in these scenarios. The evolution is similar in all three population.

**Supplemental Figure 9.** Evolution of the median PrEP coverage over the course of the Secondary Scenarios. In these scenarios, the simulation run for 10 years with each scenario starting without PrEP initially and the stopping rate stays fixed over time. The "PrEP Coverage Calibration Point" indicate where the PrEP coverage was assessed for calibration purposes.

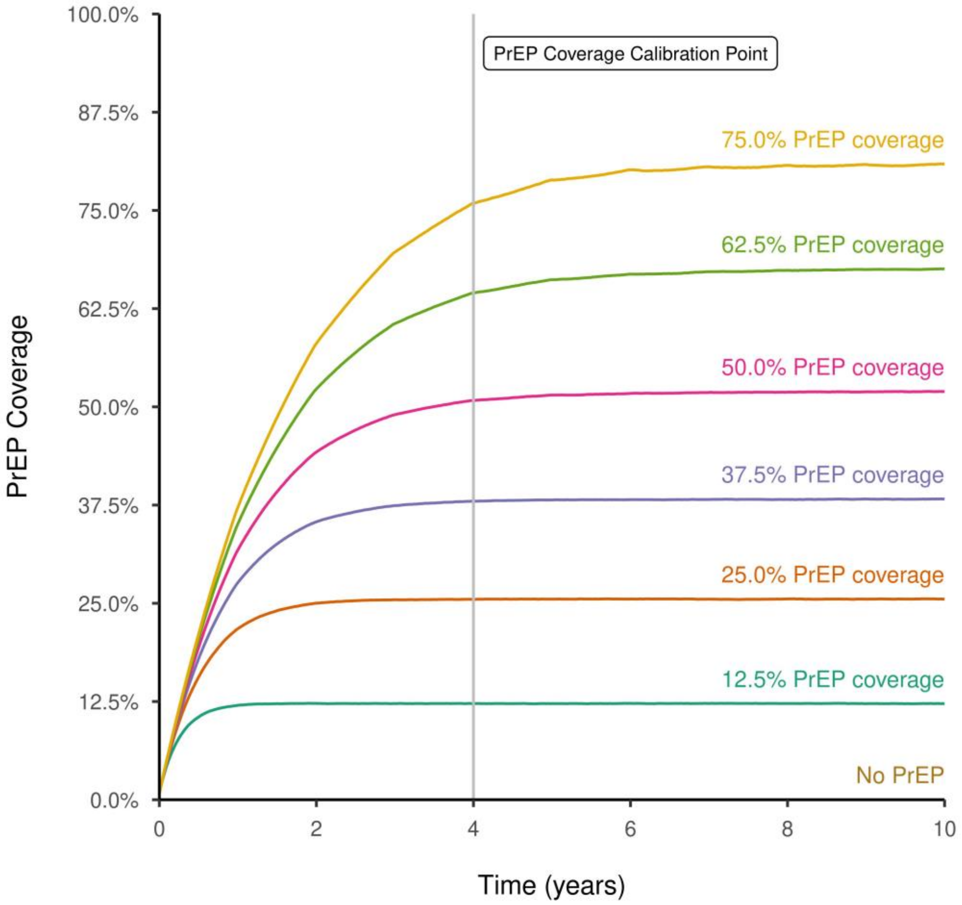

#### 11 SUPPLEMENTAL RESULTS

**Suplemental Figure 10.** Schematic representation of an HIV epidemic showing how a similar effect of PrEP (a 50% reduction of infections) result in a different overall reduction of the infectious population depending on the rate of dropping out of ART. Panels A1 and A2 represent a population with Low ART coverage and panels B1 and B2 represent a population with High ART coverage. We represented three groups: HIV-susceptible individuals (S); HIV-infected not treated (I); and HIV-infected treated by ART (Tx). We considered two time points: before PrEP (Panels A1 and B1) and after PrEP (Panel A2 and B2). Transitions from S to I were set to be 100 infections before PrEP and 50 after, reflecting the primary prevention benefit of PrEP. Transitions from I to Tx were 200 in the Low ART population and 150 in the Hight ART population, reflecting the smaller size of the I population and a similar probability of starting treatment. Transitions from Tx to I were 100 in the Low ART population and 50 in the High ART population to represent the lower rate of stopping treatment. These values ensured the stability of the I population in both cases before PrEP. After PrEP was introduced (Panels A2 and B2), the flow from S to I was halved. In the Low ART case, it reduced the total flow into I by 25% (from 200 to 150). In the High ART case, the reduction was 33% (from 150 to 100). Considering that the incidence is a function of the prevalence, we can then understand why the relative reduction will be higher in the High ART population. Supplemental Tables 16 to 18 present the flows in and out of these subpopulations over time in our simulations.

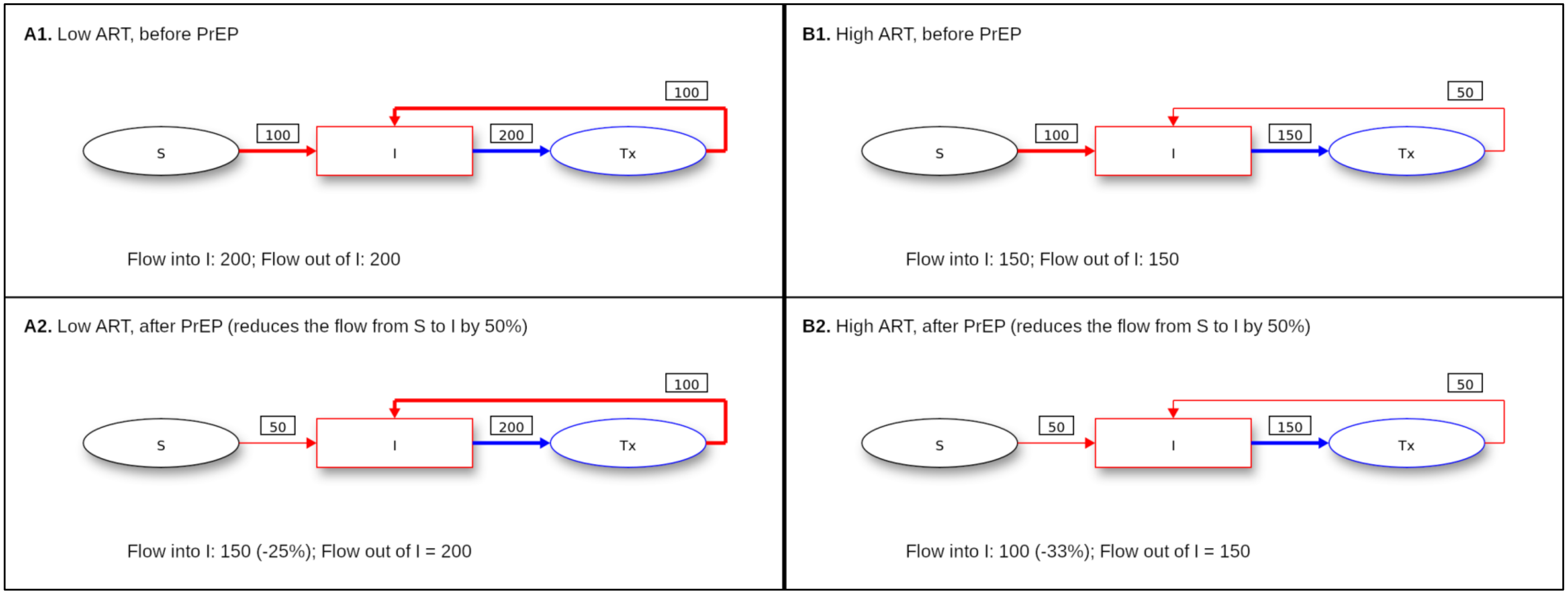

| States | | Flows | |
|---|---|---|---|
| S | HIV-susceptible (not infected) | S → I | new infections |
| I | HIV-infectious | I → Tx | Tx: ART initiation |
| TX | HIV-non-infectious Treated by ART | Tx → I | ART dropouts |

**Supplemental Table 16.** Changes in Flow from Susceptible to Infected[1] Across Secondary Scenarios.

| Scenario | | Low ART (51.2 % viral suppression) median (95% SI[2]) | Empirical ART (60% viral suppression) median (95% SI[1]) | High ART (72.9% viral suppression) median (95% SI[1]) |
|---|---|---|---|---|
| **PrEP target** | **Time** | | | |
| no PrEP (reference) | Year 0 | 0.0% (0.0%, 0.0%) | 0.0% (0.0%, 0.0%) | 0.0% (0.0%, 0.0%) |
| | Year 2 | -4.5% (-44.3%, 65.7%) | -4.0% (-55.3%, 78.6%) | -0.4% (-64.5%, 143.4%) |
| | Year 6 | -5.9% (-47.0%, 56.1%) | -1.8% (-49.3%, 86.3%) | -6.7% (-66.9%, 151.7%) |
| | Year 10 | -5.1% (-43.3%, 62.6%) | -2.0% (-50.7%, 88.8%) | -8.1% (-70.4%, 172.2%) |
| 50.0% | Year 0 | 0.0% (0.0%, 0.0%) | 0.0% (0.0%, 0.0%) | 0.0% (0.0%, 0.0%) |
| | Year 2 | -44.0% (-69.6%, 9.8%) | -47.3% (-79.3%, 5.2%) | -55.0% (-91.0%, 24.1%) |
| | Year 6 | -52.6% (-75.7%, -17.7%) | -58.6% (-84.2%, -4.8%) | -74.1% (-100.0%, -21.8%) |
| | Year 10 | -59.0% (-78.8%, -18.7%) | -63.2% (-87.3%, -22.1%) | -82.0% (-100.0%, -40.7%) |
| 75.0% | Year 0 | 0.0% (0.0%, 0.0%) | 0.0% (0.0%, 0.0%) | 0.0% (0.0%, 0.0%) |
| | Year 2 | -55.7% (-76.7%, -15.8%) | -60.0% (-85.1%, -14.5%) | -68.2% (-95.1%, -9.1%) |
| | Year 6 | -71.9% (-88.1%, -48.3%) | -78.4% (-93.0%, -47.3%) | -87.1% (-100.0%, -54.9%) |
| | Year 10 | -77.8% (-90.8%, -55.8%) | -84.3% (-97.1%, -60.0%) | -93.2% (-100.0%, -68.6%) |

1 Percentage of the susceptible that get infected, 3 months rolling mean. The changes are relative to the median value of the no PrEP Scenario by year for each ART baseline population

2 Simulation Interval.

**Supplemental Table 17.** Changes in Flow from Infectious to Treated[1] Across Secondary Scenarios.

| Scenario | | Low ART (51.2 % viral suppression) median (95% SI[2]) | Empirical ART (60% viral suppression) median (95% SI[1]) | High ART (72.9% viral suppression) median (95% SI[1]) |
|---|---|---|---|---|
| **PrEP target** | **Time** | | | |
| no PrEP (reference) | Year 0 | 0.0% (0.0%, 0.0%) | 0.0% (0.0%, 0.0%) | 0.0% (0.0%, 0.0%) |
| | Year 2 | -8.4% (-30.7%, 21.7%) | 2.0% (-35.5%, 56.5%) | -15.3% (-55.1%, 54.7%) |
| | Year 6 | -8.1% (-31.1%, 20.3%) | 4.8% (-33.5%, 61.1%) | -14.0% (-57.1%, 56.1%) |
| | Year 10 | -7.4% (-30.3%, 20.5%) | 5.8% (-29.0%, 57.0%) | -14.5% (-57.4%, 56.8%) |
| 50.0% | Year 0 | 0.0% (0.0%, 0.0%) | 0.0% (0.0%, 0.0%) | 0.0% (0.0%, 0.0%) |
| | Year 2 | -8.9% (-31.2%, 19.8%) | -0.5% (-38.5%, 51.1%) | -2.6% (-57.7%, 72.2%) |
| | Year 6 | -14.0% (-37.0%, 13.2%) | -8.4% (-41.9%, 50.1%) | -3.5% (-79.8%, 135.1%) |
| | Year 10 | -13.3% (-39.1%, 14.6%) | -7.6% (-47.1%, 43.2%) | -3.0% (-100.0%, 159.8%) |
| 75.0% | Year 0 | 0.0% (0.0%, 0.0%) | 0.0% (0.0%, 0.0%) | 0.0% (0.0%, 0.0%) |
| | Year 2 | -8.1% (-33.2%, 20.0%) | -1.5% (-37.2%, 62.7%) | -1.6% (-58.4%, 75.0%) |
| | Year 6 | -15.7% (-40.0%, 9.7%) | -12.1% (-49.2%, 33.4%) | -3.3% (-100.0%, 184.6%) |
| | Year 10 | -17.8% (-43.3%, 12.0%) | -14.7% (-50.7%, 35.1%) | -0.3% (-100.0%, 308.1%) |

1 Percentage of the infected non treated that initiate treatment, 3 months rolling mean. The changes are relative to the median value of the no PrEP Scenario by year for each ART baseline population

2 Simulation Interval.

**Supplemental Table 18.** Changes in Flow from Treated to Infectious[1] Across Secondary Scenarios.

| Scenario | | Low ART (51.2 % viral suppression) median (95% SI[2]) | Empirical ART (60% viral suppression) median (95% SI[1]) | High ART (72.9% viral suppression) median (95% SI[1]) |
|---|---|---|---|---|
| **PrEP target** | **Time** | | | |
| no PrEP (reference) | Year 0 | 0.0% (0.0%, 0.0%) | 0.0% (0.0%, 0.0%) | - [3] |
| | Year 2 | -0.5% (-27.5%, 40.5%) | -0.8% (-44.9%, 79.1%) | |
| | Year 6 | -0.1% (-30.9%, 46.0%) | 0.2% (-49.0%, 83.4%) | |
| | Year 10 | -0.6% (-31.8%, 45.9%) | -2.1% (-47.7%, 79.1%) | |
| 50.0% | Year 0 | 0.0% (0.0%, 0.0%) | 0.0% (0.0%, 0.0%) | - [3] |
| | Year 2 | -0.4% (-29.2%, 40.9%) | -3.1% (-43.0%, 70.9%) | |
| | Year 6 | 0.6% (-32.2%, 43.8%) | -1.7% (-44.5%, 74.9%) | |
| | Year 10 | 0.0% (-29.9%, 43.2%) | -1.7% (-47.6%, 74.7%) | |
| 75.0% | Year 0 | 0.0% (0.0%, 0.0%) | 0.0% (0.0%, 0.0%) | - [3] |
| | Year 2 | 0.0% (-28.4%, 45.3%) | 0.6% (-45.2%, 77.4%) | |
| | Year 6 | -1.0% (-29.8%, 42.5%) | -0.7% (-45.4%, 82.4%) | |
| | Year 10 | 0.3% (-33.7%, 50.6%) | 0.6% (-51.3%, 91.2%) | |

1 Percentage of the infected that are treated dropping out of treatment, 3 months rolling mean. The changes are relative to the median value of the no PrEP Scenario by year for each ART baseline population

2 Simulation Interval.

3 In the High ART population, no one drops out of treatment

**Supplemental Table 19.** Reduction of HIV Incidence per 100 PYAR After 5 and 10 Years of Intervention Across Secondary Scenarios.

| Scenario | **Low ART (51.2 % viral suppression)** HIV Incidence reduction[2] median (95% SI[1]) | | **Empirical ART (60% viral suppression)** HIV Incidence reduction[2] median (95% SI[1]) | | **High ART (72.9% viral suppression)** HIV Incidence reduction[2] median (95% SI[1]) | |
|---|---|---|---|---|---|---|
| | Year 5 | Year 10 | Year 5 | Year 10 | Year 5 | Year 10 |
| **PrEP target** | | | | | | |
| no PrEP (reference) | -1% (-26%, 20%) | -2% (-25%, 19%) | 6% (-24%, 31%) | 9% (-22%, 33%) | -2% (-62%, 45%) | -2% (-69%, 41%) |
| 12.5% | 12% (-10%, 33%) | 12% (-11%, 32%) | 21% (-5%, 41%) | 24% (0%, 47%) | 19% (-26%, 56%) | 25% (-29%, 62%) |
| 25.0% (SFO level) | 27% (9%, 45%) | 29% (11%, 46%) | 35% (14%, 53%) | 40% (18%, 59%) | 39% (5%, 68%) | 47% (3%, 74%) |
| 37.5% | 40% (24%, 53%) | 42% (26%, 55%) | 48% (29%, 64%) | 53% (34%, 68%) | 55% (26%, 76%) | 61% (31%, 80%) |
| 50.0% | 51% (36%, 62%) | 54% (42%, 68%) | 59% (44%, 71%) | 65% (51%, 76%) | 66% (43%, 83%) | 72% (49%, 89%) |
| 62.5% | 62% (50%, 73%) | 66% (56%, 76%) | 69% (57%, 79%) | 75% (64%, 84%) | 76% (58%, 89%) | 83% (65%, 94%) |
| 75.0% | 69% (59%, 78%) | 74% (65%, 82%) | 76% (65%, 84%) | 81% (73%, 89%) | 83% (68%, 92%) | 87% (76%, 96%) |

[1] Simulation Interval.

[2] Reduction of HIV incidence rate per 100 person-years at risk compared to the first year of the simulated intervention in the no PrEP group.

**Supplemental Table 20.** HIV Outcomes Associated with Increasing Levels of PrEP Coverage, Stratified by the Proportion of Virally Suppressed Among the Infected. For Each ART Baseline (Low, Reference, High) PIA and NNT Are Calculated Using the no PrEP Scenario of the Baseline.

| | **Low ART (51.2 % viral suppression)** | | | | **Empirical ART (60% viral suppression)** | | | | **High ART (72.9% viral suppression)** | | | |
|---|---|---|---|---|---|---|---|---|---|---|---|---|
| **Scenario** | HIV Incidence[2] | NIA[3] | PIA[4] | NNT[5] | HIV Incidence[2] | NIA[3] | PIA[4] | NNT[5] | HIV Incidence[2] | NIA[3] | PIA[4] | NNT[5] |
| | *median (95% SI¹)* | *median (95% SI¹)* | *median (95% SI¹)* | *median (95% SI¹)* | *median (95% SI¹)* | *median (95% SI¹)* | *median (95% SI¹)* | *median (95% SI¹)* | *median (95% SI¹)* | *median (95% SI¹)* | *median (95% SI¹)* | *median (95% SI¹)* |
| **PrEP target** | | | | | | | | | | | | |
| no PrEP (reference) | 2.07 (1.32, 2.97) | - | - | - | 1.33 (0.78, 2.13) | - | - | - | 0.59 (0.23, 1.21) | - | - | - |
| 12.5% | 1.73 (1.00, 2.56) | 178 (67, 294) | 12% (5%, 20%) | 36 (22, 90) | 1.07 (0.58, 1.71) | 169 (46, 282) | 16% (4%, 27%) | 45 (26, 136) | 0.39 (0.15, 0.86) | 141 (9, 252) | 23% (1%, 42%) | 64 (35, 435) |
| 25.0% (SFO level) | 1.41 (0.86, 2.18) | 362 (264, 477) | 25% (18%, 33%) | 36 (27, 49) | 0.80 (0.39, 1.30) | 326 (217, 419) | 31% (20%, 39%) | 48 (37, 71) | 0.27 (0.08, 0.62) | 251 (154, 331) | 42% (25%, 55%) | 75 (57, 120) |
| 37.5% | 1.14 (0.65, 1.76) | 516 (425, 594) | 35% (29%, 41%) | 37 (33, 44) | 0.62 (0.26, 1.11) | 449 (370, 529) | 42% (35%, 50%) | 51 (43, 61) | 0.19 (0.04, 0.46) | 326 (250, 392) | 54% (41%, 65%) | 85 (71, 109) |
| 50.0% | 0.88 (0.49, 1.45) | 664 (580, 738) | 46% (40%, 51%) | 38 (35, 43) | 0.47 (0.21, 0.83) | 557 (490, 628) | 52% (46%, 59%) | 54 (48, 61) | 0.11 (0.00, 0.34) | 386 (321, 437) | 64% (53%, 72%) | 94 (83, 113) |
| 62.5% | 0.63 (0.29, 1.02) | 798 (729, 860) | 55% (50%, 59%) | 40 (37, 43) | 0.30 (0.09, 0.60) | 656 (596, 712) | 62% (56%, 67%) | 57 (53, 62) | 0.08 (0.00, 0.23) | 425 (381, 469) | 70% (63%, 78%) | 106 (96, 117) |
| 75.0% | 0.48 (0.23, 0.77) | 892 (834, 953) | 61% (57%, 66%) | 41 (38, 43) | 0.21 (0.08, 0.43) | 720 (674, 765) | 68% (64%, 72%) | 60 (56, 64) | 0.04 (0.00, 0.15) | 455 (413, 488) | 75% (68%, 81%) | 113 (106, 124) |

1 Simulation Interval.

2 HIV incidence rate per 100 person-years at risk in the year 10 of the simulated intervention.

3 Percent of infections averted over the 10-year intervention relative to the reference model.

4 Number needed to treat to avert one infection over the 10-year intervention relative to the reference model.

5 Number of infections averted over the 10-year intervention relative to the reference model.

**Supplemental Figure 10**. Number of cumulative infections averted over 10 years of simulation for the secondary scenarios. Each color represents a baseline ART coverage, and the x axis shows the different levels of PrEP coverage, from 0% to 75%.

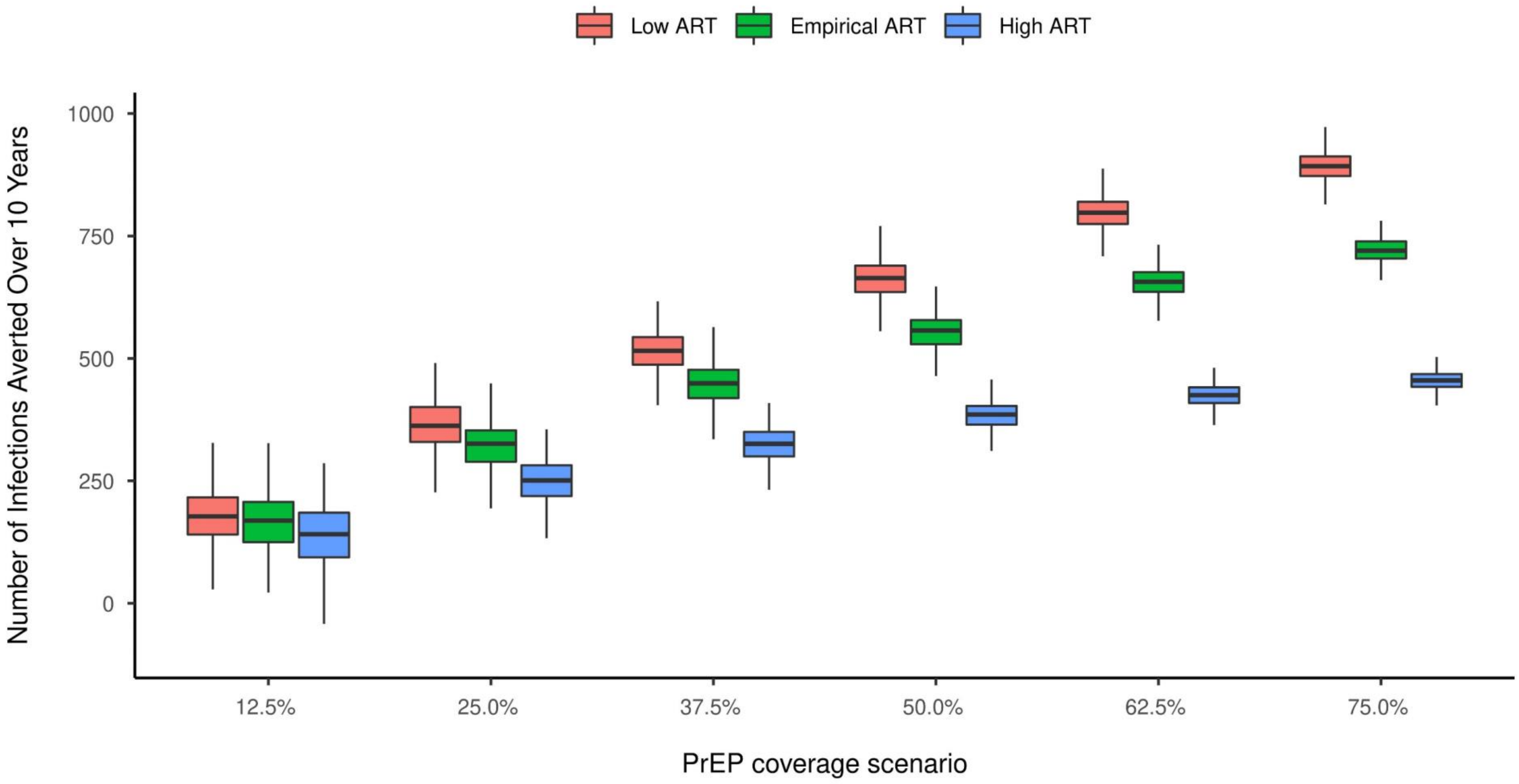

**Supplemental Figure 11**. Percent of infections averted over 10 years of simulation for the secondary scenarios. Each color represents a baseline ART coverage, and the x axis shows the different levels of PrEP coverage, from 0% to 75%.

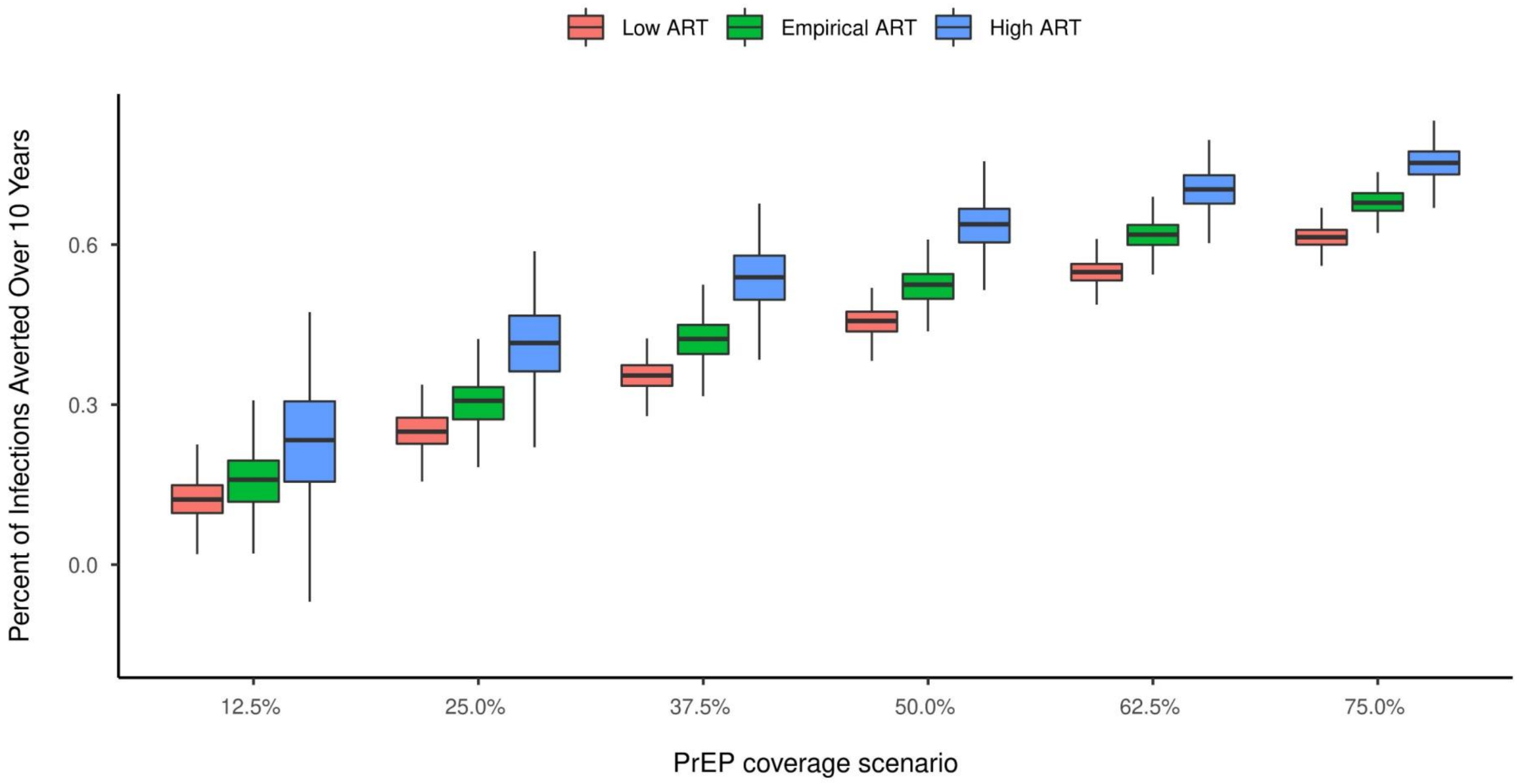

**Supplemental Figure 12** Number needed treat to avert one infection over 10 years of simulation for the secondary scenarios. Each color represents a baseline ART coverage, and the x axis shows the different levels of PrEP coverage, from 0% to 75%..

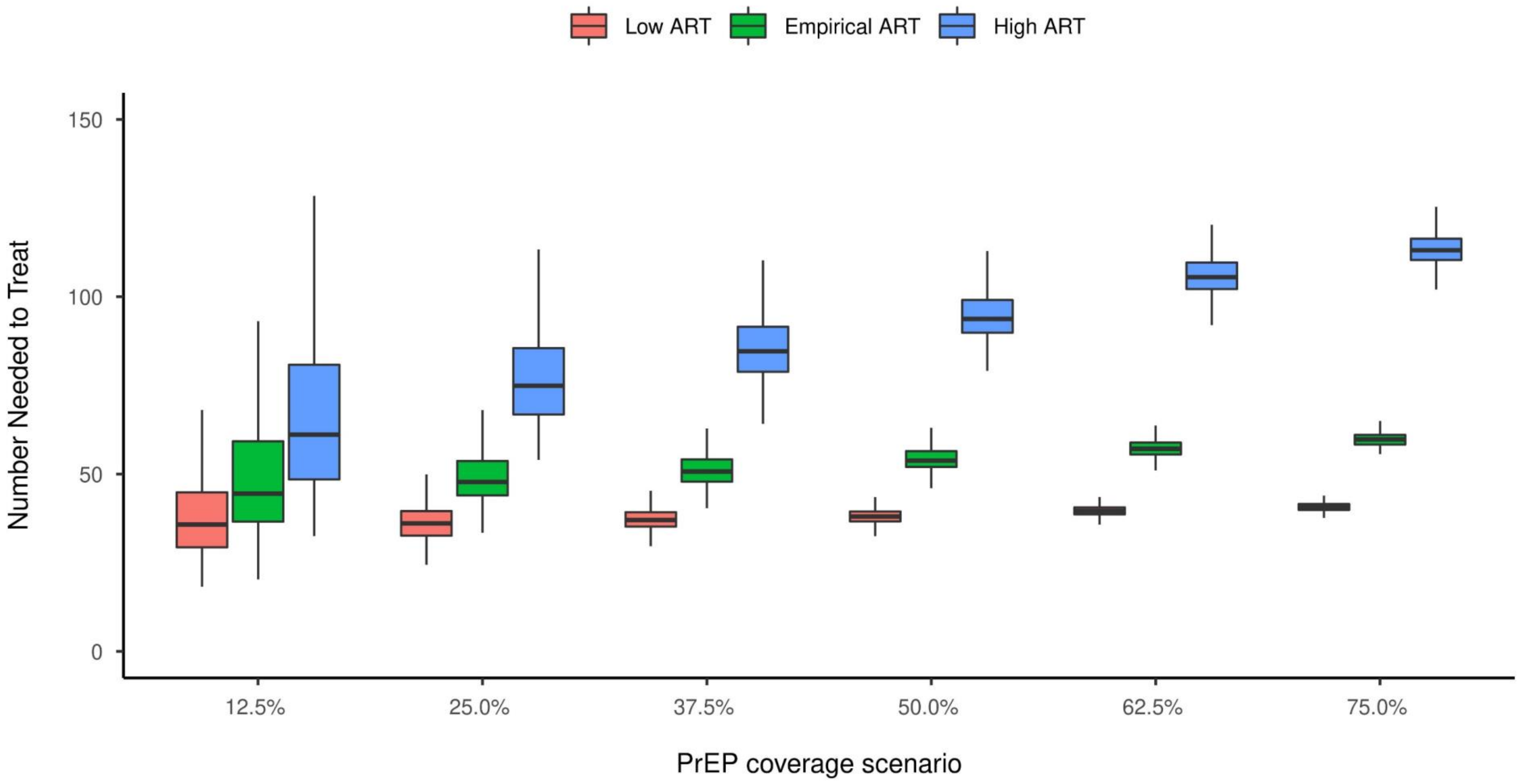

**Supplemental Figure 13.** Evolution of the median HIV prevalence over the course of the Secondary Scenarios. This shows the absolute reduction of HIV prevalence over 10 years with each additional increase in PrEP coverage, in every baseline ART population

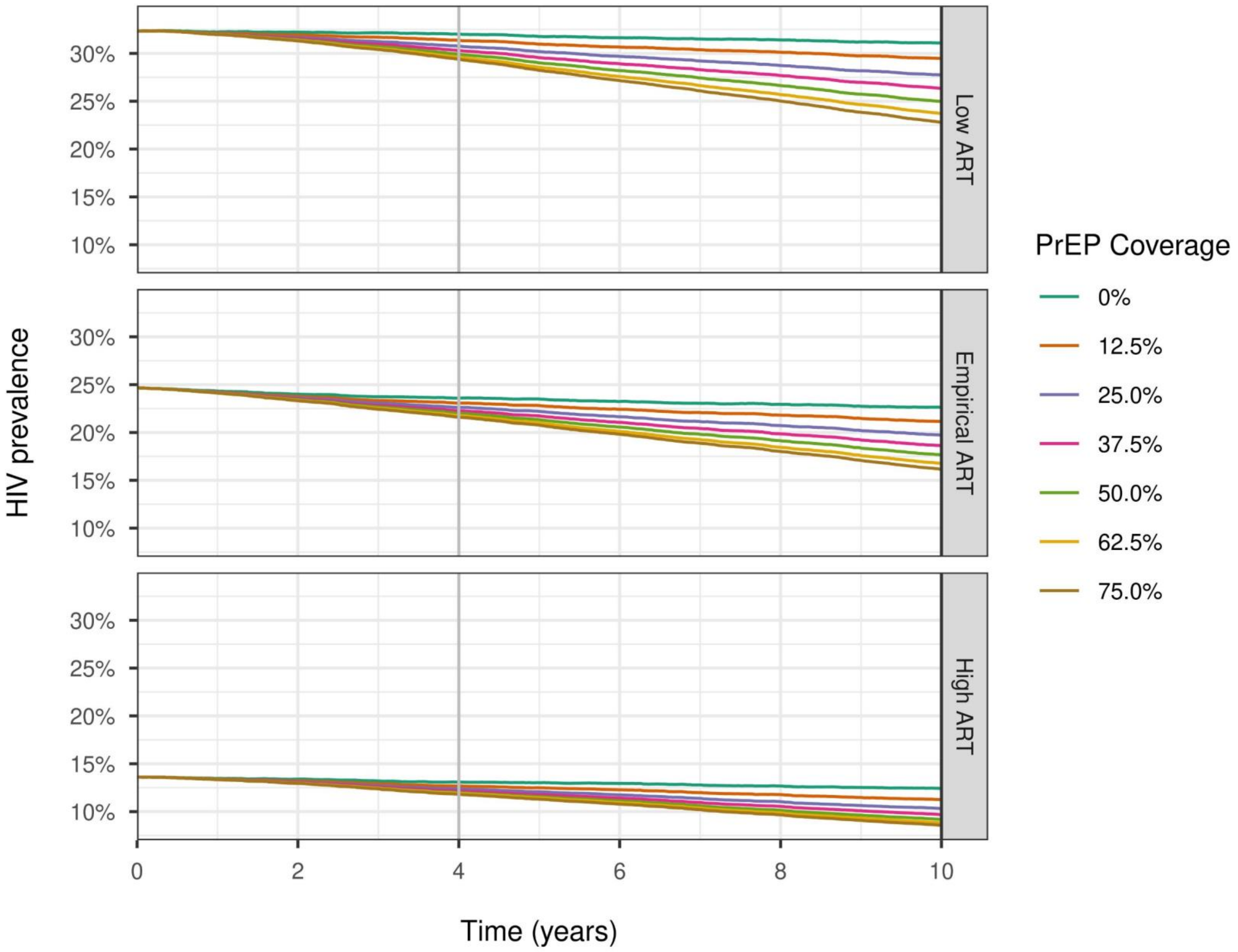

**Supplemental Figure 14.** Evolution of the median HIV incidence over the course of the Secondary Scenarios. This shows the reduction of HIV incidence over 10 years with each additional increase in PrEP coverage, in every baseline ART population.

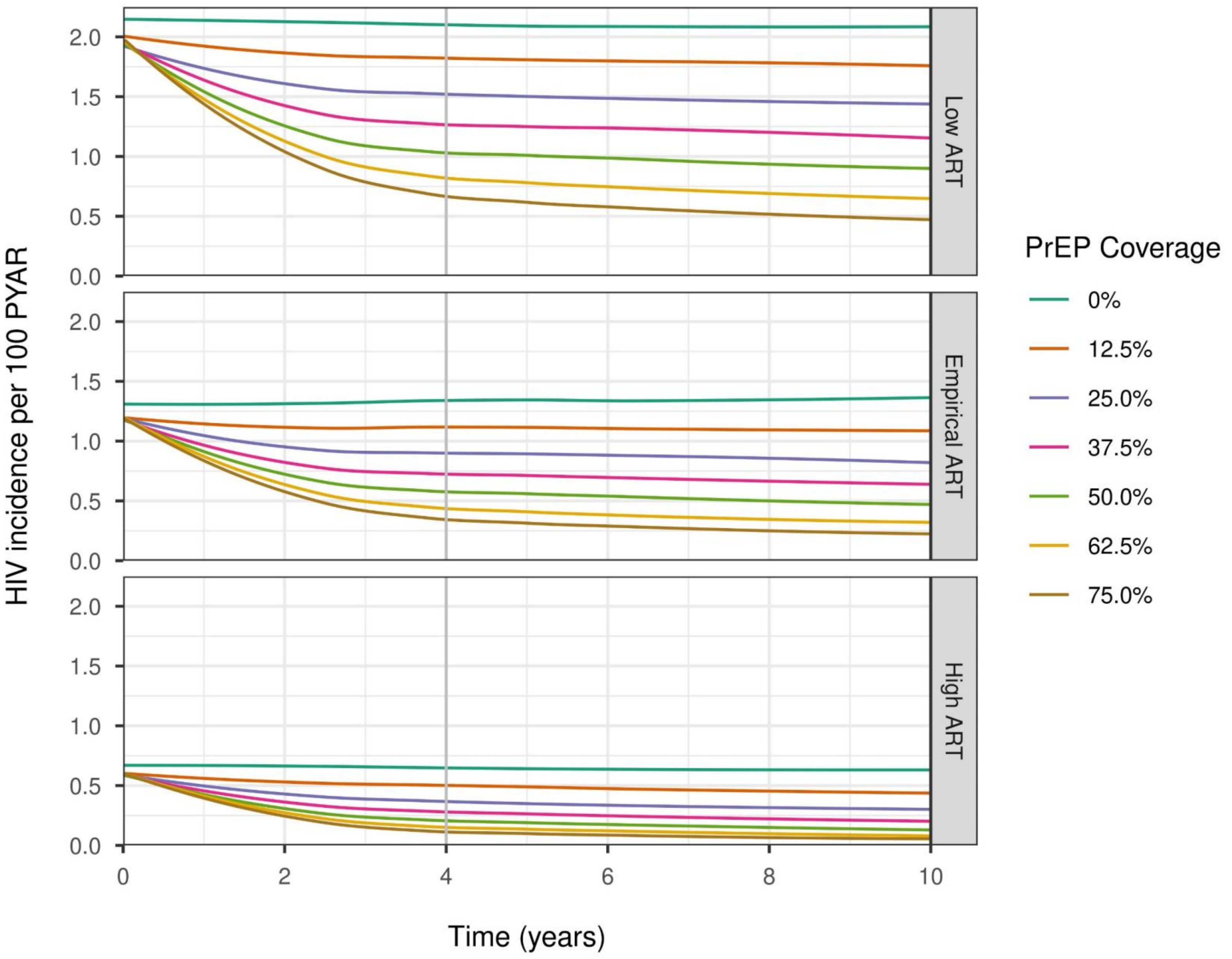
